# Wavelet-derived indices of cardiac vagal and vasomotor oscillations as a new tool to better understand autonomic imbalance in post-COVID-19 dysautonomia

**DOI:** 10.64898/2026.11.14.26367993

**Authors:** I Romero Fragoso, A Sáinz-Jiménez, T Reyes-Cruz, E Chiquete, KM Tamez-Torres, J Sifuentes-Osornio, R Fossion, B Estañol, JJ Aceves-Buendía

## Abstract

**Background:** When aiming to quantify autonomic imbalance, conventional heart rate variability (HRV) analysis generally reduces each posture to a single spectral value derived from interbeat intervals (IBI) alone. This analysis based only in IBI obscures transient events during blood pressure changes. We propose two wavelet-derived indices that quantify cardiac vagal and vasomotor sympathetic oscillations separately and over time. Since autonomic imbalance is directly related to baroreflex sensitivity (BRS), we evaluated them against an estimate of baroreflex sensitivity in post-COVID-19 patients.

**Methods:** In this study, 121 participants (32 healthy controls and 89 patients with persistent post-COVID-19 autonomic symptoms) underwent an active standing test with continuous finger blood pressure. The continuous wavelet transform with generalized Morse wavelets yielded time-varying high-frequency power of IBI and low-frequency power of systolic blood pressure (SBP). Two new indices were proposed: Index *I* combined the magnitude and variability of HF IBI power (aimed to quantify parasympathetic activity), and the index *J* combined LF SBP power with the rate of its transient elevations (bursts), which aims to quantify sympathetic vasomotor activity; J/I was their ratio. Indices were computed for supine and orthostatic periods and compared with conventional HRV and BRS measurements.

**Results:** Index *I* correlated strongly with HF power of IBI and was lower in all patient subgroups than in controls, Index *J* had only a weak relation with LF power of IBI. On standing, *J* increased most in the subgroups with the worst baroreflex function. Orthostatic J/I was higher than in controls amongst patients, while LF/HF showed no differences. J/I was inversely correlated with BRS, most strongly when standing, and more strongly than LF/HF also. Adding *J* to *I* raised the cross-validated variance explained in orthostatic BRS substantially. In one example patient, a window during symptoms showed vasomotor oscillations that were hidden in the posture average.

**Conclusions:** Wavelet-derived indices from routine beat-to-beat recordings separate cardiac vagal modulation from sympathetic vasomotor oscillations and their transients. J/I correlates with BRS impairment more closely than LF/HF, providing a potential alternative estimate of autonomic balance, and its time-resolved visual output allows analysis of clinically relevant windows within a posture. The new proposed variables allowed an evaluation of autonomic imbalance that could be applied to different pathologies.

## 1 Introduction

Orthostatic intolerance and other manifestations of cardiovascular autonomic dysfunction are common among individuals with persistent symptoms following SARS-CoV-2 infection (Davis et al., 2023; Schoene et al., 2026). Notably, many of these patients do not meet the established diagnostic criteria for postural orthostatic tachycardia syndrome (POTS) despite experiencing clear symptoms and exhibiting abnormal hemodynamic responses to standing (Sáinz-Jiménez et al., 2026). The arterial baroreflex plays a central role in regulating these responses. Upon standing, blood pools below the diaphragm, reducing venous return and cardiac filling, while unloading arterial baroreceptors. These changes trigger compensatory responses involving cardiac vagal withdrawal and increased cardiac and vascular sympathetic activity (Jordan et al., 2023). In a previous study, we demonstrated that the graphical estimate of baroreflex sensitivity (BRS), represented by the θ angle, identifies a spectrum of baroreflex impairment in patients with post-COVID-19 conditions that is not captured by conventional diagnostic categories (Sáinz-Jiménez et al., 2026).

In clinical practice, cardiovascular autonomic function is commonly assessed using heart rate variability (HRV), including time-domain indices and stationary spectral power techniques within the low-frequency (LF, 0.04–0.15 Hz) and high-frequency (HF, 0.15–0.40 Hz) bands (Malik et al., 1996). However, three major limitations restrict the utility of this approach in the assessment of dysautonomia. First, conventional Fourier-based spectral analysis assumes signal stationarity and typically summarizes an entire recording period, such as a given postural condition, into a single value, thereby overlooking transient changes within that period (Malik et al., 1996; Pichot et al., 1999). This is particularly relevant in post-COVID-19 conditions, whose symptoms, including autonomic ones, fluctuate in severity (Zanin et al., 2023) and can follow an episodic, relapsing course (Owen et al., 2025). Second, the physiological interpretation of low-frequency (LF) power and, consequently, of the LF/HF ratio remains controversial. Although the LF/HF ratio has been proposed as an index of sympathovagal balance (Furlan et al., 2000), LF power derived from interbeat intervals (IBI) does not directly reflect cardiac sympathetic outflow and largely reflects baroreflex-mediated responses to blood pressure oscillations (Cevese et al., 2001; Goldstein et al., 2011; Mitsuyama et al., 2024). Consistent with this ambiguity, LF/HF findings in post-COVID-19 conditions have been inconsistent, ranging from increased values (Mooren et al., 2023) to no significant pooled difference from healthy controls in a recent meta-analysis (Schoene et al., 2026). Third, because these conventional HRV indices are derived exclusively from IBI, they fail to capture potentially relevant information contained in concurrent blood pressure fluctuations.

These limitations have important practical implications. During the visual inspection of beat-to-beat recordings obtained during active standing tests in our laboratory, physicians identified transient episodes in which blood pressure and heart rate (HR) increased simultaneously. Such episodes run counter to the reciprocal changes expected from an intact baroreflex, yet they are lost when each posture is summarized by a single spectral value. Time-frequency analysis offers a way to preserve this temporal information. However, the short-time Fourier transform relies on a fixed analysis window, imposing a trade-off between temporal and frequency resolution (Mallat, 1999; Mitsuyama et al., 2024). In contrast, the continuous wavelet transform (CWT) adapts its effective window to each frequency and provides a continuous time-frequency representation suited to nonstationary physiological signals (Torrence and Compo, 1998; Kralj and Lenasi, 2023). Previous studies have also shown that wavelet-based analysis of IBI can detect autonomic changes that may be overlooked or identified later by conventional Fourier-based methods (Pichot et al., 1999; Wang et al., 2021).

Previous time-frequency approaches to autonomic assessment can be broadly classified into two categories. The first tracks HRV spectral indices over time, including time-varying LF/HF ratios, but relies exclusively on IBI-derived information (Pichot et al., 1999; Chen, 2002; Lu et al., 2005; Wang et al., 2021). The second estimates time-varying BRS using cross-spectral analysis of systolic blood pressure (SBP) and IBI; these methods restrict estimates to time-frequency regions that exhibit significant coherence and, in some methods, a phase relationship consistent with baroreflex-mediated cardiac responses, thereby discarding information from regions that do not meet these criteria (Orini et al., 2012; Kazimierska et al., 2019; de Boer and Karemaker, 2019; Chao-Ecija and Dawid-Milner, 2023). Although these approaches have advanced the characterization of baroreflex dynamics, they primarily quantify the cardiac component of the reflex and they are not designed to show the sympathetic vasomotor and cardiac vagal components separately.

In particular, the vascular component, reflected in LF oscillations of SBP, which include the ∼0.1 Hz oscillations known as Mayer waves, has received comparatively less attention, despite its close association with sympathetic vasomotor activity (Julien, 2006; Brychta et al., 2007). Although the LF power of SBP is not a direct surrogate of muscle sympathetic nerve activity (MSNA) at the individual level (Ryan et al., 2011; Julien, 2020), it has been proposed as a marker of sympathetic vasomotor modulation (Furlan et al., 2000; Brychta et al., 2007).

From a systems theory perspective, HR can be viewed as a responsive variable that contributes to buffering cardiovascular perturbations, whereas arterial blood pressure is a regulated variable whose stability depends, in part, on these compensatory responses. Under healthy conditions, this framework predicts greater variability in the responsive component and relatively lower variability in the regulated variable, with this relationship potentially becoming disrupted in disease (Fossion et al., 2018). Accordingly, impaired baroreflex function may be reflected in both components of cardiovascular regulation: reduced cardiac vagal modulation, reflected in a decrease in HF IBI oscillations, and enhanced or poorly buffered LF SBP oscillations (Julien, 2006; Goldstein et al., 2011). In this study, we use the term autonomic imbalance for this combination: a predominance of sympathetic vasomotor oscillations over cardiac vagal modulation.

A tool is therefore needed that identifies and quantifies each of these components over time, using their respective effector signals (IBI and SBP), over a user-defined time window. Such indices would complement conventional stationary HRV measures and help to establish a new way to describe autonomic imbalance.

In this study, we propose two CWT-derived indices obtained from routine beat-to-beat recordings: Index ***I***, which characterizes HF IBI power and its variability as an index of cardiac vagal modulation, and the index ***J***, which characterizes LF SBP power with the rate of transient increases (bursts) in LF power as an index of vasomotor sympathetic modulation, as well as their ratio, ***J/I.*** Both indices are accompanied by scalograms and time-varying band-power curves, which allow the temporal evolution of each component to be visualized and facilitate the selection of a time window of interest for targeted analysis. As an initial evaluation of these indices, we calculated them over the supine and orthostatic periods of an active standing test in healthy controls and in post-COVID-19 patients classified according to their θ-based baroreflex phenotype (Sáinz-Jiménez et al., 2026). We hypothesized that patients with greater baroreflex impairment would exhibit lower ***I*** index values, a greater orthostatic increase in ***J***, and higher ***J/I*** ratios. We also compared **J/I** with the conventional LF/HF ratio.

## 2 Methods

### 2.1 Participants, ethics, and θ-based baroreflex phenotyping

This cross-sectional study was approved by the Research Ethics Committee at the National Institute of Medical Sciences and Nutrition Salvador Zubirán (INCMNSZ; approval NER-4340 22-24-1) and was conducted in accordance with local legislation and institutional requirements and the Declaration of Helsinki. All participants provided written informed consent.

Participants were recruited and screened following the procedures described by Sáinz-Jiménez et al. (2026). All participants were enrolled through the same recruitment process; the present cohort included 32 healthy controls and 89 patients. Patients included in the present study were individuals with persistent symptoms for at least four weeks after a mild to moderate SARS-CoV-2 infection that had not required hospitalization. Healthy controls had no known comorbidities, including diabetes or hypertension, and no known previous SARS-CoV-2 infection.

Baroreflex function was characterized with the θ angle, a graphical estimator of BRS previously developed and evaluated by our group, lower values indicate lower BRS (Sáinz-Jiménez et al., 2026). Patients were classified into three baroreflex phenotypes according to the criteria established in that study: (i) MP-S, patients meeting neither of the following criteria (n = 34); (ii) MP-Δθ>40, a decrease in θ of at least 40° from the supine to the orthostatic position (Δθ ≤ −40°; n = 19); and (iii) MP-θ<40, a θ BRS indicator below 40° in both postures (n = 36). Given the progressive reduction in baroreflex function across these phenotypes (Sáinz-Jiménez et al., 2026), MP-S, MP-Δθ>40 and MP-θ<40 are hereafter referred to as *slight*, *moderate* and *severe* baroreflex impairment, respectively.

### 2.2 Active standing protocol, recordings and preprocessing

Cardiovascular recordings were obtained during an active standing test, following consensus recommendations for orthostatic evaluation (Jordan et al., 2023). Participants remained supine, motionless and silent for 7 min, breathing spontaneously and avoiding sighs. They then stood up without assistance and remained upright for an additional 7 min under the same conditions. Symptoms reported by participants during the test were time-marked on the recording.

Continuous non-invasive blood pressure was recorded using a Finometer (Finapres Medical Systems, Enschede, The Netherlands) at 200 Hz, with the finger cuff placed on the ring finger of the left hand, as described previously (Sáinz-Jiménez et al., 2026). Beat-to-beat SBP and IBI were exported from the Finometer software with their corresponding timestamps. Because IBI was derived from successive arterial pressure pulses, it represents the pulse interval, which was used as a surrogate of the ECG-derived RR interval (Kazimierska et al., 2019).

For each participant, a 14-min recording segment spanning the postural transition was extracted, and the moment of standing was identified using the event marker. The SBP and IBI series were visually inspected, and artifacts and anomalous beats were excluded based on abnormal amplitude and discontinuities relative to neighboring beats; on average, 0.13% of beats were removed per recording, a proportion reported as recommended by the Task Force (Malik et al., 1996). The resulting gaps were not interpolated at this stage; they were bridged during the resampling step described in section 2.4.

To exclude the initial cardiovascular response to the postural change, a 60-s interval centered on the standing transition was removed. This resulted in two analysis segments of approximately 6.5 min each, one supine and one orthostatic, which were used for all measures in the study, including θ-derived BRS (Section 2.1), conventional HRV and hemodynamic variables (Section 2.3), and the wavelet-derived indices (Sections 2.4–2.5). Transient fluctuations within each postural period were retained, as their characterization is one of the objectives of the proposed method (Section 2.5.2). Posture-based segments allowed comparison with conventional measures; however, the indices can be calculated over any time window of at least 90 s (Section 2.6).

### 2.3 Conventional HRV and hemodynamic variables

Conventional HRV indices were computed using Kubios HRV Lite (version 4.2.0; Tarvainen et al., 2014) from the beat-to-beat IBI series corresponding to the supine and orthostatic analysis segments. Time-domain analysis included the root mean square of successive differences (RMSSD) and the percentage of successive intervals differing by more than 50 ms (pNN50). Frequency-domain analysis included absolute LF (0.04–0.15 Hz) and HF (0.15–0.40 Hz) power, as well as the LF/HF ratio (Malik et al., 1996). Spectral estimates were obtained using the fast Fourier transform (FFT)-based (Welch) method after resampling the IBI series at 4 Hz and applying smoothness-priors detrending (λ = 500). These indices served as reference measures for comparison with the proposed wavelet-derived indices. Specifically, HF power and RMSSD were used as conventional indices of cardiac vagal modulation, and LF power of IBI was the conventional LF index, traditionally (although controversially) interpreted as a marker of sympathetic activity.

Hemodynamic variables were obtained from the same analysis segments. Beat-to-beat SBP (mmHg), IBI (ms), and HR (bpm) were exported directly from the Finometer software and averaged separately for each segment.

For all variables, the orthostatic change was calculated as Δ*X* = *X_ort_*_ℎ*o*_− *X_sup_*, with positive values indicating an increase upon standing.

### 2.4 Wavelet time-frequency analysis

The wavelet analysis was designed to characterize the temporal evolution of two oscillatory components associated with distinct aspects of cardiovascular autonomic regulation: LF oscillations of SBP (0.04–0.15 Hz), which include the ∼0.1 Hz oscillations known as Mayer waves and are closely associated with sympathetic vasomotor modulation (Julien, 2006; Brychta et al., 2007), and HF oscillations of IBI (0.15–0.40 Hz), which predominantly reflect respiratory sinus arrhythmia and are largely mediated by cardiac vagal modulation (Malik et al., 1996).

We used the CWT, which adapts its effective temporal resolution to the frequency being analyzed and allows power to be integrated over frequency bands whose boundaries match the physiological ranges of interest (Torrence and Compo, 1998; Mallat, 1999). The choice of the CWT over the short-time Fourier transform and the discrete wavelet transform is detailed in supplementary methods (Table S1).

Because the beat-to-beat series are unevenly sampled, SBP and IBI were resampled at 4 Hz by piecewise cubic Hermite interpolation (PCHIP; Fritsch and Carlson, 1980), and each resampled series was linearly detrended. No amplitude normalization was applied to the input signals, allowing the wavelet power to retain its dependence on the magnitude of the underlying SBP and IBI oscillations; wavelet power is reported in arbitrary units (a.u.). The CWT was computed over the entire recording, and the supine and orthostatic analysis segments (Section 2.2) were extracted from the resulting time-frequency representation, avoiding an artificial signal boundary at the postural transition.

The CWT was computed in Python with the ssqueezepy package (version 0.6.5; Muradeli, 2024) using generalized Morse wavelets (Olhede and Walden, 2002; Lilly and Olhede, 2012), which have previously been used for wavelet-based baroreflex analysis (de Boer and Karemaker, 2019), with γ = 3 for both signals. The time-bandwidth product (P²) was set to 20 for SBP, favoring frequency resolution for the slower LF oscillations, and to 10 for IBI, favoring temporal resolution for the faster and more transient HF oscillations. Because the choice of wavelet and its parameters influences spectral estimates (Tzabazis et al., 2018), all settings are reported in full in Table S1.

Time-varying band power was obtained by integrating the squared magnitude of the CWT coefficients over the band of interest with the trapezoidal rule, after after coefficients affected by edge effects, i.e., at frequencies below the cone-of-influence (COI) boundary, were set to zero (Torrence and Compo, 1998):

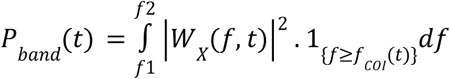

where *W _X_*(*f*, *t*) is the complex wavelet coefficient of signal X (IBI for HF analysis and SBP for LF analysis) at frequency f and time t, and the integration limits [f1, f2] are 0.04–0.15 Hz for SBP (*P*(*t*)*_LF_*) and 0.15–0.40 Hz for IBI (*P*(*t*)*_HF_*).

The resulting scalograms and band-power curves are illustrated for a representative control and patient in Figure 1 (A, B IBI recordings for a control and a patient, respectively, C, D the CWT IBI-derived scalograms for the same subjects) and Figure 2 (A, B SBP recordings for a control and a patient, respectively, C, D the CWT SBP-derived scalograms for the same subjects).

**Figure 1.**
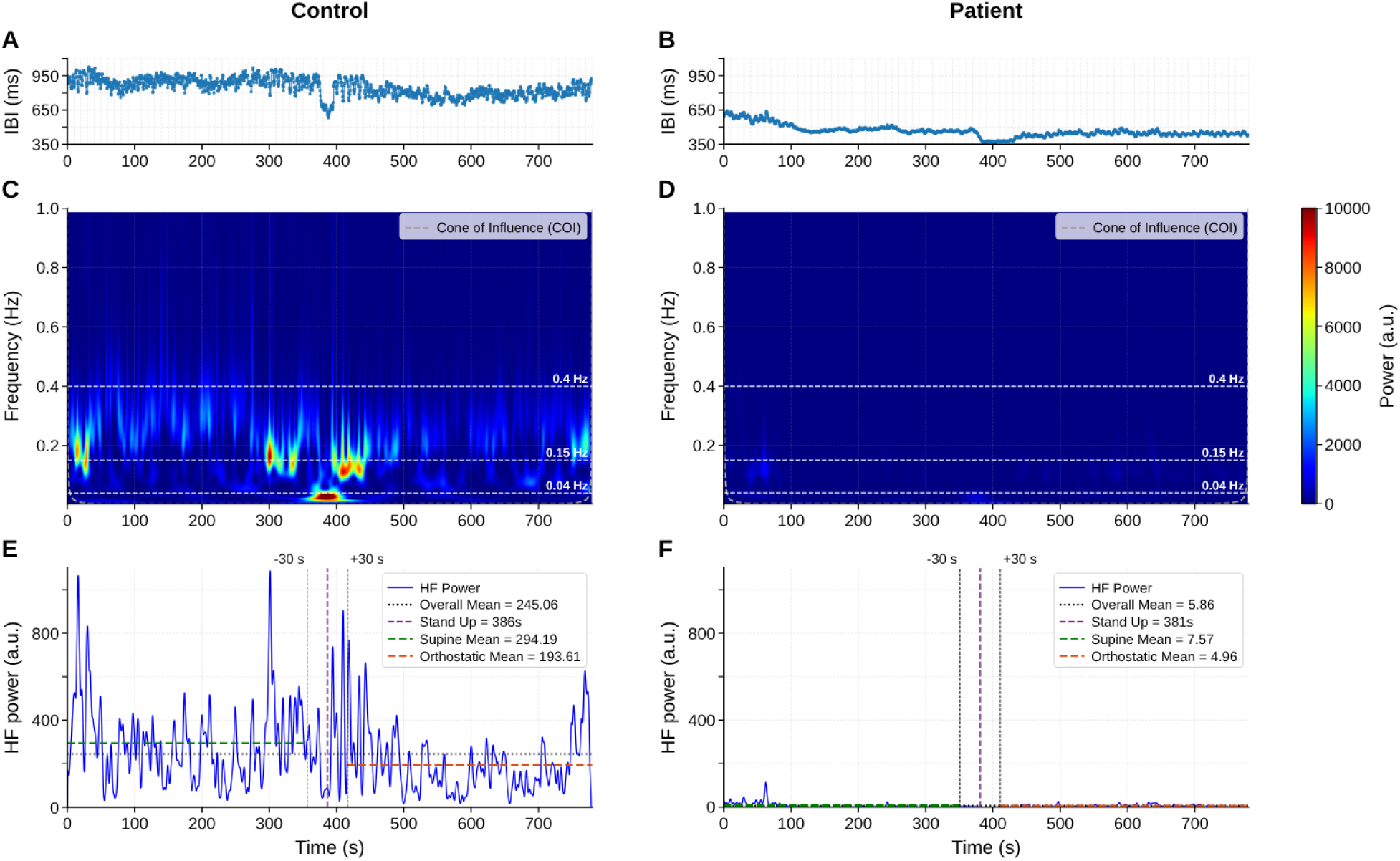
Wavelet-based analysis of HF power from IBI for the derivation of index. *I*. The left column shows a representative control, whereas the right column shows a patient from the severe subgroup. (A, B) Beat-to-beat interbeat interval (IBI) during the active standing test. (C, D) Wavelet power scalograms of IBI, with white dashed lines indicating the frequency boundaries (0.04, 0.15, and 0.40 Hz) and the grey dashed contour marking the cone of influence (COI). (E, F) Time-varying high-frequency (HF) power,, extracted within the 0.15–0.40 Hz band. The purple dashed line indicates standing onset, black dashed lines delimit the excluded ±30-s posture change interval, and horizontal lines represent mean HF power over the entire recording (black dotted) and during supine (green) and orthostatic (orange) periods. The control shows relatively high supine HF power that decreases upon standing, whereas the patient exhibits markedly reduced HF power in both postures.

**Figure 2.**
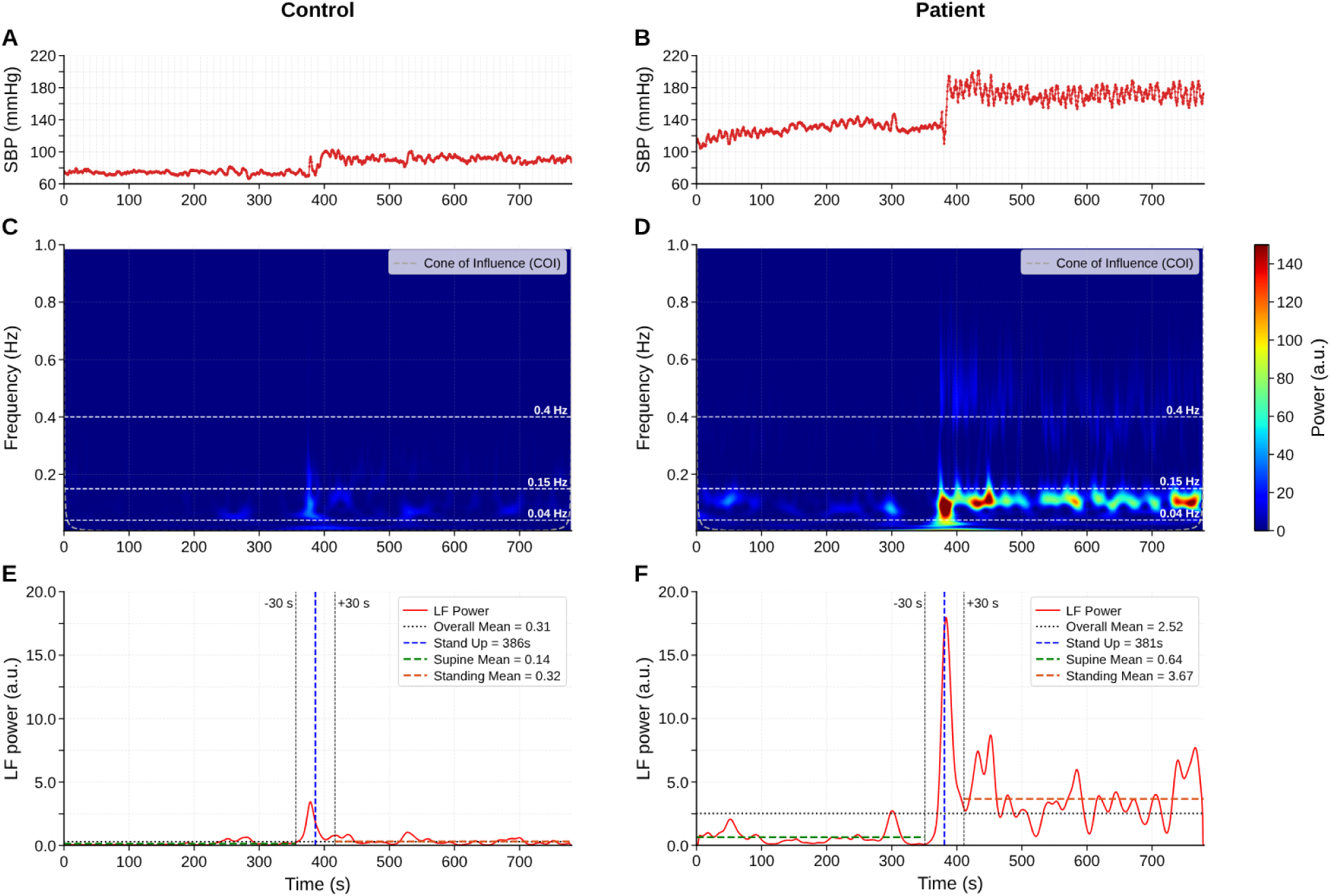
Wavelet-based analysis of LF power from systolic blood pressure (SBP) for the derivation of index. **J.** The left column shows the same representative control as in Figure 1, whereas the right column shows the same patient from the severe subgroup. (A, B) Beat-to-beat systolic blood pressure (SBP) during the active standing test. The step change reflects the transition from supine to standing. (C, D) Wavelet power scalograms of SBP, with white dashed lines indicating the frequency boundaries (0.04, 0.15, and 0.40 Hz). (E, F) Time-varying low-frequency (LF) power of SBP, extracted within the 0.04–0.15 Hz band. The blue dashed line indicates standing onset, black dashed lines delimit the excluded ±30-s posture change interval, and horizontal lines represent mean LF power over the entire recording (black dotted) and during supine (green) and orthostatic (orange) periods. In the control, SBP LF power remains relatively low throughout the test (supine mean: 0.14 a.u.; orthostatic mean: 0.32 a.u.). In contrast, the patient exhibits a marked increase in oscillations after standing (from 0.64 to 3.67 a.u.), with repeated transient elevations.

### 2.5 Proposed indices

For each analysis segment s (supine or orthostatic), with boundaries *t_a_* and *t_b_*, two indices were derived from the band-power curves described in section 2.4: Index ***I*** from *P*(*t*)*_HF_* of IBI and index ***J*** from *P*(*t*)*_LF_* of SBP. Each combines a component describing the overall magnitude of the oscillation with a component describing its temporal dynamics within the segment.

#### 2.5.1 Index ***I***: HF oscillations of IBI

The magnitude component was the area under the HF power curve normalized by segment duration in minutes:

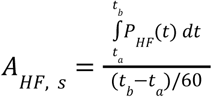

The dynamic component was the standard deviation of *P_HF_* (*t*) within the segment, σ*_HF_*_, *s*_, which describes how much HF power fluctuated over time. Index ***I*** was defined as:

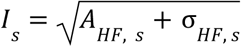

The square root compresses the wide inter-individual range of wavelet power, with σ*_HF_*_, *s*_ adding information on its temporal fluctuation. Figure 1E, F shows *P_HF_*(*t*), the excluded posture-transition window and the segment means for a representative control and patient.

#### 2.5.2 Index ***J***: LF oscillations of SBP and transient bursts

The magnitude component was defined as for index ***I***:

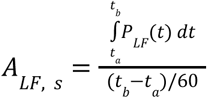

The dynamic component was designed to capture transient increases in LF SBP power (bursts) occurring within a given posture, representing the type of within-posture episode that motivated this work (Section 1). Bursts were detected in *P_LF_*(*t*) within each segment against a time-varying baseline, m(t), obtained with a centered 90-s moving average. A burst began when *P_LF_*(*t*) exceeded m(t) + 1 a.u. and ended when it fell back below m(t), and the maximum of each completed event was taken as its peak. Because an event had to return to baseline before a new one could start, each oscillatory episode was counted once.

The burst rate was calculated as:

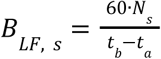

Where *N_s_* is the number of bursts in the segment, expressed in bursts per minute.

The detection threshold was defined empirically, in consensus with the clinical team of the Neuroinflammation and Dysautonomia Laboratory (INCMNSZ). The team reviewed the LF SBP power curves obtained from this cohort against the symptoms that participants reported and that were time-marked during the recordings. In most patients, symptomatic episodes coincided with excursions of *P _LF_* (*t*) above approximately 2 a.u., and increases greater than 1 a.u. above the local baseline were judged clinically meaningful. Because wavelet power was not normalized, this threshold is specific to the present acquisition and processing settings.

Index ***J*** was defined as:

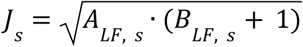

The additive constant prevents ***J*** from becoming zero when no bursts are detected. Figure 2E, F shows *P _LF_* (*t*) for a representative control and patient.

#### 2.5.3 Combined index ***J/I***

The combined index was calculated for each segment as

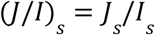

without further normalization. Higher ***J/I*** values indicate predominance of LF SBP oscillations relative to HF IBI oscillations, whether from a higher index J, a lower index I, or both. This is the pattern expected when impaired baroreflex function reduces cardiac vagal buffering of vasomotor oscillations, i.e., the autonomic imbalance defined in Section 1.. Unlike LF/HF, which is computed entirely from IBI, the numerator and denominator of ***J/I*** come from different effector signals: the vasculature (SBP) and the heart (IBI). ***J/I*** contrasts different bands from different signals, LF for SBP and HF for IBI, so that each reflects its predominant autonomic influence.

### 2.6 Visual output and window of interest analysis

In addition to the indices, the method generates scalograms of the IBI and SBP signals, together with time-varying band-power curves *P_HF_* (*t*) and *P _LF_* (*t*) (Figures 1 and 2). These outputs are intended to support visual inspection by clinicians. They show how HF IBI and LF SBP power evolve over time within each postural period, allowing changes in these components to be examined in relation to relevant symptoms during the recordings and concurrent changes in SBP and HR. After a window of interest has been identified, the indices ***I***, ***J***, and ***J/I*** can be calculated for that window using the definitions provided in Section 2.5, with *t_a_* and *t_b_* set to the corresponding window limits. The minimum window length is 90 s. In the present study, window-of-interest analysis was applied only to the illustrative case in Section 3.8 (Figure 8); all statistical analyses used the posture-based segments. This corresponds to the length of the moving-average baseline used for burst detection and to more than three cycles of the slowest LF oscillation (one cycle lasts 25 s at 0.04 Hz). Because the CWT is computed over the entire recording before any window is extracted (section 2.4), windows that lie away from the beginning and end of the recording are not affected by edge effects.

### 2.7 Statistical analysis

Statistical analyses were performed in Python (version 3.13.14) using NumPy (2.2.6), SciPy (1.16.0), pandas (2.3.1), scikit-learn (1.6.1) and statsmodels (0.15.0). Continuous variables are reported as median (interquartile range) and categorical variables as number and percentage n(%). Sex distribution across the four groups was compared with Pearson’s χ² test.

Segment-related variables and their orthostatic changes were compared across the four groups with the Kruskal–Wallis test. Each patient subgroup was then compared with controls, regardless of the Kruskal–Wallis result, using two-sided Mann–Whitney U tests, with Bonferroni correction for the three comparisons per variable. Because the patient subgroups were defined from θ-derived BRS, between-group differences in θ are reported descriptively.

Associations were assessed at the individual level (n = 121) using two-sided Spearman rank correlations (ρ). These analyses included the associations of indices ***I*** and ***J*** with conventional HRV indices (HF power, RMSSD and LF power of IBI) and of ***I***, ***J***, ***J/I*** and LF/HF with θ-derived BRS, each computed separately for the supine and orthostatic periods. Fitted curves with 95% confidence bands were overlaid to visualize nonlinear relationships: power-law curves in Figure 3 (least squares on log-transformed variables) and exponential growth (index ***I***) or exponential decay to a plateau (***J***, ***J/I*** and LF/HF; nonlinear least squares) in Figure 5. They were descriptive only and did not affect the correlation analyses.

**Figure 3.**
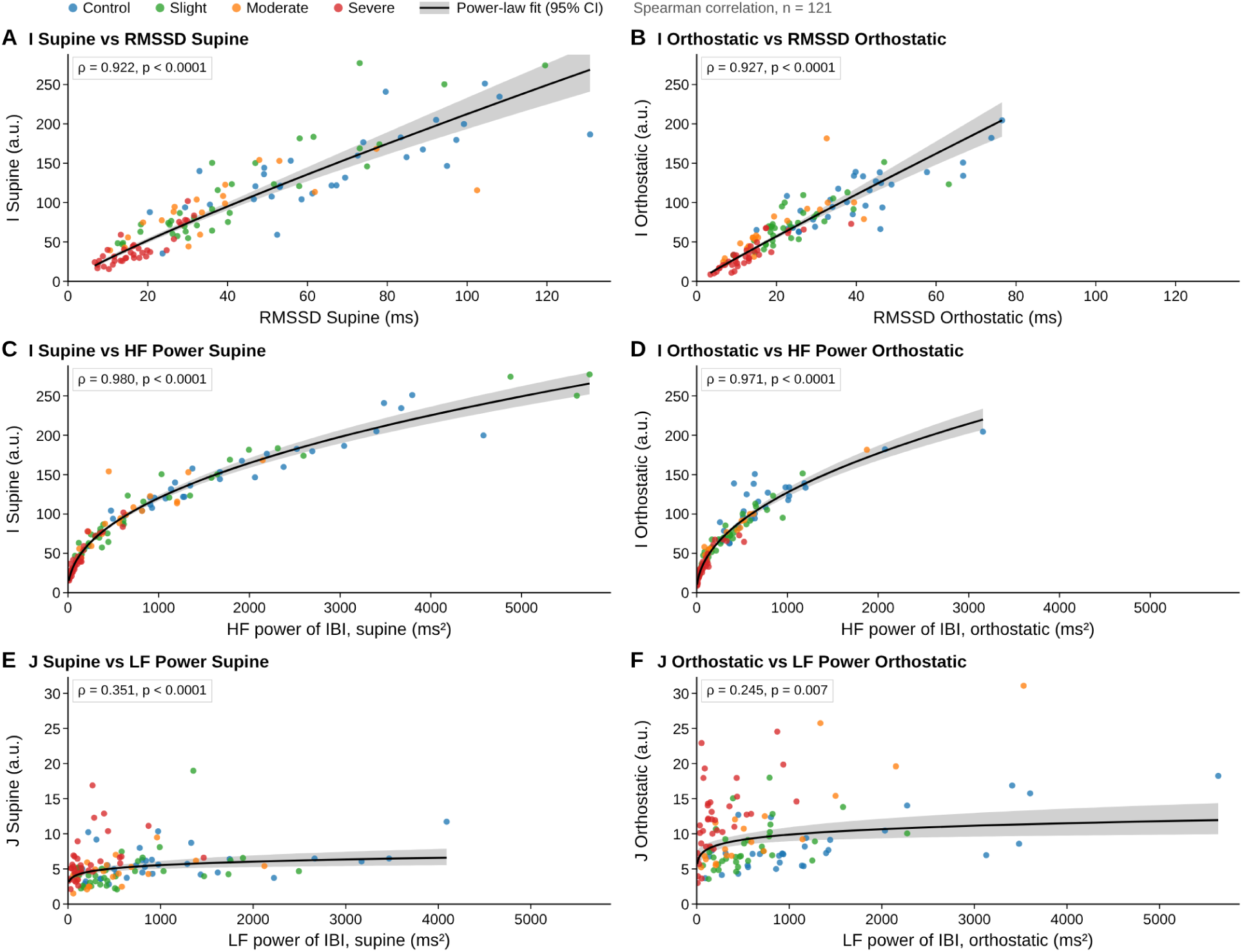
Relationships of wavelet-derived indices with conventional HRV measures. (A,B) *I* versus RMSSD, (C,D) *I* versus HF power of IBI, and (E, F) *J* versus LF power of IBI, during supine (left) and orthostatic (right) periods. Each point represents one participant (n = 121), with colors indicating study group (control, slight, moderate, and severe). Black curves represent power-law fits with 95% confidence bands and are shown as visual guides. Spearman’s ρ and p values correspond to two-sided correlation tests. *I* and *J* are wavelet-derived indices, whereas RMSSD and LF and HF power of IBI (ms²) were calculated using Kubios HRV from the same analysis segments.

To assess whether ***J/I*** was more strongly associated with θ-derived BRS than LF/HF, participants were resampled with replacement 10,000 times (random seed 20261101). In each resample, the difference between the absolute Spearman correlations, |ρ(***J/I***, θ)| − |ρ(LF/HF, θ)|, was computed. Its 95% confidence interval was obtained from the 2.5th and 97.5th percentiles of the bootstrap distribution, separately for each posture. A confidence interval excluding zero was taken as evidence of a stronger association for ***J/I***.

To evaluate whether ***J*** contributed information on orthostatic θ-derived BRS beyond ***I*** index, two nested linear regression models were fitted with *ln*_(_θ*_ort_*_ℎ*o*)_ as the dependent variable: a reduced model with *ln*_(_*I_ort_*_ℎ*o*)_ and a full model with *ln*_(_*I_ort_*_ℎ*o*)_ and *ln*_(_*J_ort_*_ℎ*o*)_. Logarithms were used to linearize the relationships. Predictive performance was evaluated with 100 repetitions of shuffled 10-fold cross-validation and summarized as the mean cross-validated coefficient of determination (R²) and root mean squared error (RMSE). Because the dependent variable was log-transformed, exp(RMSE) was calculated to express prediction error as a multiplicative factor on the original scale. The incremental contribution of ***J*** was quantified as ΔR² = R²(***I*** + ***J***) − R²(**I**) in each repetition and summarized by its mean and 2.5th–97.5th percentiles. Model coefficients were obtained by fitting each model to the complete sample (n = 121).

The classification performance of conventional HRV indices (LF, HF, and LF/HF ratio) and the proposed indices (I, J, and J/I) was evaluated using θ-defined baroreflex impairment as the reference condition, with thresholds of θ < 68.3° during the supine period and θ < 40.3° during orthostatism. Sensitivity, specificity, positive and negative predictive values (PPV and NPV), positive and negative likelihood ratios (LR+ and LR−), and overall accuracy were calculated. Optimal cutoff values were determined by maximizing the Youden index (sensitivity + specificity – 1). Accuracy was calculated as the proportion of correctly classified participants. All performance estimates were reported with 95% confidence intervals (Wilson score for proportions, logarithmic method for likelihood ratios, Newcombe hybrid score for the Youden index).

Statistical significance was set at p < 0.05, using Bonferroni-adjusted p values for the group comparisons.

## 3 Results

### 3.1 Participants and hemodynamic response to standing

A total of 121 participants were included in the analysis: 32 healthy controls and 89 patients with persistent post-COVID-19 autonomic symptoms. According to the θ-based classification (Section 2.1), 34 patients were assigned to the slight subgroup, 19 to the moderate subgroup and 36 to the severe subgroup (Table 1). Women accounted for 56.2% of the healthy controls and 77.5% of the patients, with proportions ranging from 75.0 to 79.4% across the three patient subgroups. This distribution is consistent with the female predominance reported in post-COVID-19 conditions (Davis et al., 2023; Shahbaz et al., 2025). Sex distribution did not differ significantly across the four groups (χ²(3) = 5.47, p = 0.141).

**Table 1.**
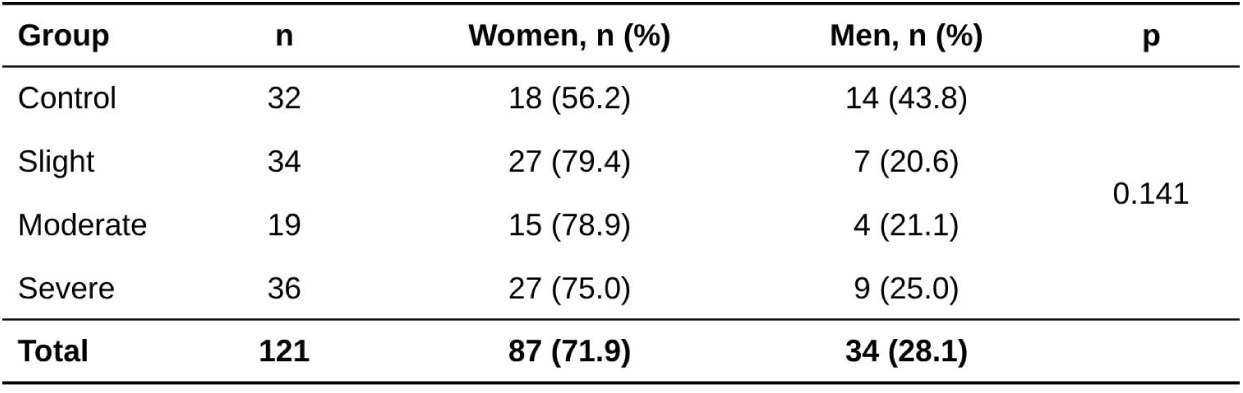
Study population and sex distribution across the four study groups. Participants were classified into four groups: healthy controls and patients with slight (MP-S; neither criterion met), moderate (MP-Δθ>40; decrease in θ of at least 40° during orthostasis) and severe (MP-θ<40; θ <40° in both supine and orthostatic conditions) baroreflex impairment. Data are presented as the number of participants and sex distribution, n (%). Sex distribution did not differ significantly across groups (Pearson’s χ²(3) = 5.47, p = 0.141).

| **Group** | **n** | **Women, n (%)** | **Men, n (%)** | **p** |
|---|---|---|---|---|
| Control | 32 | 18 (56.2) | 14 (43.8) | 0.141 |
| Slight | 34 | 27 (79.4) | 7 (20.6) |  |
| Moderate | 19 | 15 (78.9) | 4 (21.1) |  |
| Severe | 36 | 27 (75.0) | 9 (25.0) |  |
| **Total** | **121** | **87 (71.9)** | **34 (28.1)** |  |

Supine SBP was comparable across groups (p = 0.326; Table 2). During orthostasis, SBP differed across groups (p = 0.019) and was higher in the severe subgroup than in controls (median 121.1 vs. 107.0 mmHg; adjusted p = 0.010). The orthostatic change in SBP, however, did not differ across groups (p = 0.240). All three patient subgroups had a higher HR than controls in both postures (supine medians: 69.4–78.5 vs. 62.4 bpm; orthostatic: 84.8–91.2 vs. 74.8 bpm; all adjusted p ≤ 0.005).

**Table 2.**
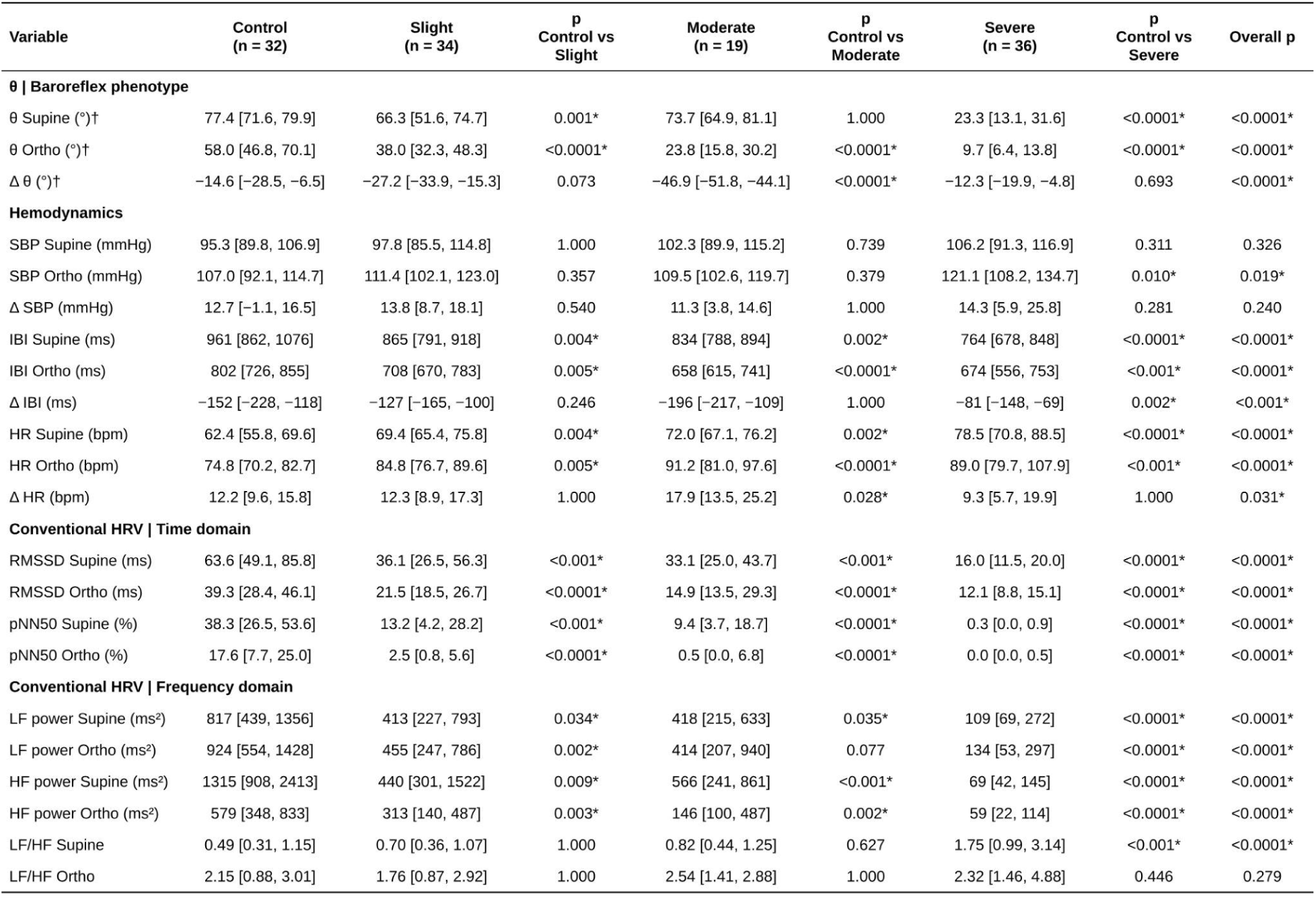
θ-based phenotype variables, hemodynamic measures, and conventional HRV indices across the four study groups. Values are presented as median [interquartile range (IQR), 25th–75th percentile]. Patient subgroups were compared with controls using two-sided Mann–Whitney U tests with Bonferroni correction for the three comparisons per variable. Overall group differences were assessed using the Kruskal–Wallis test. Bold values marked with an asterisk indicate p < 0.05.

The magnitude of the orthostatic cardiac response differed across groups (p=0.031). The increase in HR upon standing was greater in the moderate subgroup than in controls (median ΔHR 17.9 vs. 12.2 bpm; adjusted p = 0.028), whereas in the severe subgroup it did not differ significantly from controls (9.3 vs. 12.2 bpm; adjusted p = 1.000).

### 3.2 Conventional HRV indices are reduced across phenotypes, whereas LF/HF does not distinguish them during orthostasis

Time-domain HRV indices were lower in all three patient subgroups than in healthy controls in both postures (Table 2). Supine RMSSD was 36.1 ms in the slight subgroup, 33.1 ms in the moderate and 16.0 ms in the severe, compared with 63.6 ms in healthy controls (all adjusted p < 0.001). During orthostasis, the corresponding values were 21.5, 14.9 and 12.1 ms vs. 39.3 ms (all adjusted p < 0.0001). pNN50 showed the same pattern and was close to zero in the severe subgroup in both postures (median 0.3% supine and 0.0% orthostatic, vs. 38.3% and 17.6% in healthy controls; both adjusted p < 0.0001).

Frequency-domain indices showed a broadly similar pattern. The HF power of IBI was lower in all three patient subgroups than in controls in both postures (all adjusted p < 0.01), with the lowest values in the severe subgroup (supine 69 vs. 1,315 ms² in controls; orthostatic 59 vs. 579 ms²). LF power of IBI was also reduced in all three subgroups in the supine position. During orthostasis, LF power was lower in the slight and severe subgroups (adjusted p = 0.002 and p < 0.0001, respectively) but not in the moderate subgroup (adjusted p = 0.077). For both indices, median values showed a graded decrease, from intermediate values in the slight and moderate subgroups to the lowest values in the severe subgroup, which also had the lowest θ-derived BRS in both postures.

Given the ongoing controversy over the ability of the LF/HF ratio to quantify sympathovagal balance (Goldstein et al., 2011; Clemson et al., 2022), we examined the LF/HF ratio separately, as it is the conventional comparator for **J/I**.

The LF/HF ratio showed a different pattern. In the supine position, LF/HF differed across groups (p < 0.0001), but only the severe subgroup differed significantly from healthy controls (median 1.75 vs. 0.49; adjusted p < 0.001). Upon standing, median LF/HF increased in every group, as expected for the orthostatic shift of spectral HRV (Malik et al., 1996; Furlan et al., 2000), and the groups converged to similar values (healthy controls 2.15; patient subgroups 1.76–2.54; overall p = 0.279). The severe subgroup no longer differed from healthy controls (adjusted p = 0.446), even though it had the lowest orthostatic θ-derived BRS.

Overall, conventional HRV indices showed a graded reduction in cardiac variability across phenotypes, particularly in the time-domain and absolute frequency-domain measures. However, the LF/HF ratio, conventionally interpreted as an index of sympathovagal balance, did not distinguish the phenotypes during orthostasis, which is the condition in which θ showed baroreflex impairment in all three subgroups. Since conventional HRV indices are derived exclusively from IBI, we next examined how the proposed indices, ***I*** and ***J***, relate to these measures and whether they provide complementary information.

### 3.3 *I* closely tracks conventional vagal HRV indices, whereas index *J* carries new SBP information

The two proposed indices showed distinct patterns of association with conventional HRV measures (Figure 3). Index ***I*** was strongly correlated with the HF power of IBI (ρ = 0.980 supine and ρ= 0.971 orthostatic; both p < 0.0001; Figure 3C,D). It was also strongly correlated with RMSSD (ρ = 0.922 supine and ρ = 0.927 orthostatic; both p < 0.0001; Figure 3A, B). These associations were virtually unchanged between postures and extended across the full range of values, from controls to the most impaired participants of the severe subgroup. The relationship between index ***I*** and HF power was concave, consistent with the square-root formulation of ***I***, whereas its relationship with RMSSD was approximately linear. Given that ***I*** and HF power both quantify HF IBI power, this close agreement mainly confirms that index ***I*** preserves the information of the conventional vagal index.

In contrast, ***J*** showed only weak correlations with LF power of IBI, the conventional HRV measure in the same frequency band (ρ = 0.351, p < 0.0001 supine; ρ = 0.245, p = 0.007 orthostatic; Figures 3E, F). The two shared at most 12% of their rank variance (ρ² ≤ 0.12), and the association weakened further with standing. The dissociation was most evident in the severe subgroup during orthostasis. This subgroup combined the highest orthostatic ***J*** of all groups (median 11.99 vs. 7.16 in controls; Table 3) with the lowest LF power of IBI (134 vs. 924 ms²; Table 2). ***J*** captured LF SBP dynamics that were largely not reflected in IBI. Having established what each index represents, we next compared indices ***I***, ***J*** and ***J/I*** across the θ-defined phenotypes in each posture.

**Table 3.** Proposed wavelet-derived indices (I, J, and J/I) across the four study groups. Values are presented as median [interquartile range (IQR), 25th–75th percentile]. Two-sided Mann–Whitney U tests for each patient subgroup versus controls, with Bonferroni correction for the three comparisons per variable, and Kruskal–Wallis tests across the four groups. Bold values marked with an asterisk indicate p < 0.05. Index *I* and index *J* (a.u.) were derived from the CWT band-power curves over the same supine and orthostatic segments used for conventional HRV analysis. J/I is dimensionless. Abbreviations: HF, high frequency; LF, low frequency.

| **Variable** | **Control<br>(n = 32)** | **Slight<br>(n = 34)** | **p<br>Control vs<br>Slight** | **Moderate<br>(n = 19)** | **p<br>Control vs<br>Moderate** | **Severe<br>(n = 36)** | **p<br>Control vs<br>Severe** | **Overall p** |
|---|---|---|---|---|---|---|---|---|
| **I \| HF-related index** |  |  |  |  |  |  |  |  |
| I Supine (a.u.) | 138.0 [110.6, 177.3] | 83.8 [65.7, 149.2] | 0.020* | 94.6 [67.0, 114.5] | 0.002* | 35.0 [26.2, 44.9] | <0.0001* | <0.0001* |
| I Ortho (a.u.) | 102.0 [76.2, 133.5] | 69.4 [52.3, 86.5] | <0.001* | 55.6 [46.3, 86.8] | <0.001* | 29.7 [20.6, 39.4] | <0.0001* | <0.0001* |
| Δ I (a.u.) | −42.8 [−60.7, −18.1] | −19.0 [−69.1, −6.5] | 1.000 | −33.1 [−51.8, −13.4] | 1.000 | −7.1 [−11.4, −0.2] | <0.001* | <0.001* |
| **J \| LF-related index** |  |  |  |  |  |  |  |  |
| J Supine (a.u.) | 5.53 [4.28, 6.42] | 4.07 [3.11, 5.13] | 0.033* | 4.33 [2.95, 5.47] | 0.138 | 5.23 [4.45, 6.66] | 1.000 | 0.011* |
| J Ortho (a.u.) | 7.16 [5.80, 9.19] | 7.16 [6.11, 9.53] | 1.000 | 8.83 [6.85, 12.27] | 0.491 | 11.99 [8.52, 14.35] | 0.002* | <0.001* |
| Δ J (a.u.) | 2.15 [0.46, 4.37] | 3.21 [2.04, 4.94] | 0.348 | 4.31 [3.13, 7.08] | 0.015* | 5.86 [2.51, 8.35] | 0.009* | 0.002* |
| **J/I \| Ratio** |  |  |  |  |  |  |  |  |
| J/I Supine | 0.036 [0.028, 0.053] | 0.047 [0.027, 0.065] | 1.000 | 0.052 [0.038, 0.061] | 0.440 | 0.143 [0.106, 0.207] | <0.0001* | <0.0001* |
| J/I Ortho | 0.082 [0.057, 0.109] | 0.120 [0.076, 0.146] | 0.005* | 0.154 [0.121, 0.212] | <0.0001* | 0.379 [0.277, 0.550] | <0.0001* | <0.0001* |
| Δ J/I | 0.039 [0.015, 0.068] | 0.066 [0.036, 0.097] | 0.018* | 0.110 [0.078, 0.159] | <0.0001* | 0.207 [0.109, 0.331] | <0.0001* | <0.0001* |

### 3.4 *I* is reduced across all phenotypes, whereas group differences in *J* follow the phenotype ordering under orthostatic stress

Index ***I*** was lower in all three patient subgroups than in healthy controls in both postures (Table 3; Figure 4A, B), consistent with its close correspondence with the HF power of IBI. Supine medians were 83.8 in the slight subgroup, 94.6 in the moderate one and 35.0 in the severe, compared with 138.0 in controls (adjusted p = 0.020, 0.002 and < 0.0001, respectively). During orthostasis, the corresponding median values were 69.4, 55.6 and 29.7 vs. 102.0 (all adjusted p < 0.001). Median ***I*** decreased with standing in every group (Figure 4C). The decrease was smallest in the severe subgroup (median Δ***I*** −7.1 vs. −42.8 in controls; adjusted p < 0.001), consistent with the already low supine values in this subgroup. Δ***I*** did not differ from controls in the slight or moderate subgroups (adjusted p = 1.000 for both).

**Figure 4.**
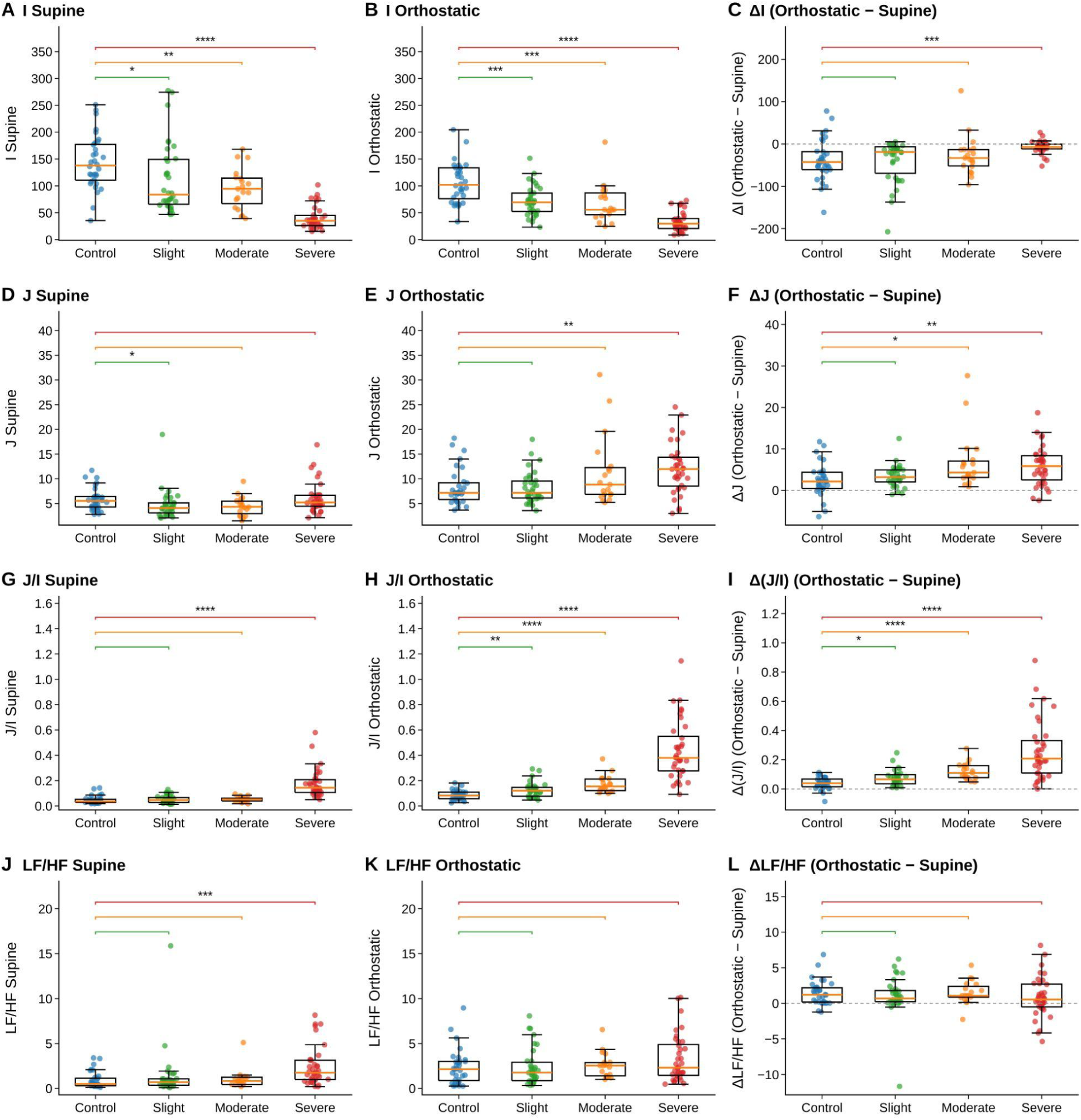
Distribution of wavelet-derived indices and conventional LF/HF ratio across study groups and postural conditions. Rows show (A–C) I, (D–F) J, (G–I) J/I, and (J–L) the conventional LF/HF ratio of IBI. Columns represent the supine period (left), orthostatic period (middle), and orthostatic change (right; Δ = orthostatic − supine). The dashed horizontal line in the right column indicates zero change. Boxes represent the median (orange line) and interquartile range (IQR), whiskers extend to 1.5 × IQR, and individual points represent participants (n = 121). Brackets indicate comparisons between each patient subgroup and controls (two-sided Mann–Whitney U tests with Bonferroni correction for three comparisons): *p < 0.05; **p < 0.01; ***p < 0.001; ****p < 0.0001. *I* and *J* are expressed in arbitrary units (a.u.), whereas J/I and LF/HF are dimensionless.

Index ***J*** showed a posture-dependent pattern. In the supine position, differences between groups were small (overall p = 0.011; Figure 4D). The only significant pairwise difference was a lower ***J*** in the slight subgroup than in healthy controls (4.07 vs. 5.53; adjusted p = 0.033), which did not follow the ordering of θ-derived BRS values across the patient subgroups. Upon standing, median ***J*** increased in all groups, but the magnitude of the increase varied. Orthostatic ***J*** values followed an increase pattern from controls and the slight subgroup (both 7.16) to the moderate subgroup (8.83) and the severe one (11.99). This ordering was inversely aligned with the median orthostatic values of θ across the groups (58.0°, 38.0°, 23.8° and 9.7°; Table 2).

The difference from healthy controls was statistically significant for the severe subgroup (adjusted p = 0.002; Figure 4E). The within-subject change showed the same gradient (Figure 4F): median Δ***J*** was 2.15 in controls, 3.21 in the slight subgroup, 4.31 in the moderate one and 5.86 in the severe subgroup and was larger than in controls for the moderate and severe subgroups (adjusted p = 0.015 and 0.009). Thus, in the moderate subgroup, the orthostatic response of ***J*** was therefore detectable as a change from each participant’s own supine value, even though absolute orthostatic index ***J*** did not differ from controls.

Combining the two indices gave the clearest separation between groups. In the supine position, ***J/I*** was elevated only in the severe subgroup (median 0.143 vs. 0.036 in controls; adjusted p < 0.0001; Figure 4G). During orthostasis, ***J/I*** was higher than in controls in all three patient subgroups: 0.120 in the slight, 0.154 in the moderate one and 0.379 in the severe subgroup, vs. 0.082 in controls (adjusted p = 0.005, < 0.0001 and < 0.0001, respectively; Figure 4H). Δ(***J/I***) likewise differed from controls in all three subgroups (0.066, 0.110 and 0.207 vs. 0.039; adjusted p = 0.018, < 0.0001 and < 0.0001; Figure 4I). Both orthostatic ***J/I*** and Δ(***J/I***) followed the ordering of orthostatic θ-derived BRS.

The conventional LF/HF ratio, shown on the same layout for comparison, differed from controls only in the severe subgroup at rest (Figure 4J). Neither orthostatic LF/HF nor its orthostatic change differed from controls in any subgroup (Figure 4K, L).

Orthostatic stress reduced ***I*** and increased ***J*** in every group, but only the increase in ***J*** scaled with the θ-defined phenotype. As a result, ***J/I*** separated every patient subgroup from controls during orthostasis, whereas LF/HF separated none. Given that the group-level patterns followed the ordering of orthostatic θ-derived BRS, we next examined whether the relationship held at the individual level, treating θ-derived BRS as a continuous variable.

### 3.5 *J/I* is strongly associated with θ-based BRS during orthostasis

When examined across participants, the proposed indices showed distinct associations with θ-derived BRS in the two postures (Figure 5). Index ***I*** was positively associated with θ in both the supine (ρ = 0.790, p < 0.0001) and orthostatic periods (ρ = 0.792, p < 0.0001; Figure 5A, B), and the strength of the association was virtually identical between postures. In contrast, ***J*** was not significantly correlated with θ in the supine position (ρ = −0.129, p = 0.160; Figure 5C). During orthostasis, however, ***J*** showed a moderate inverse association with θ (ρ = −0.401, p < 0.0001; Figure 5D), with the highest ***J*** values concentrated among participants with orthostatic θ values below approximately 20°.

**Figure 5.**
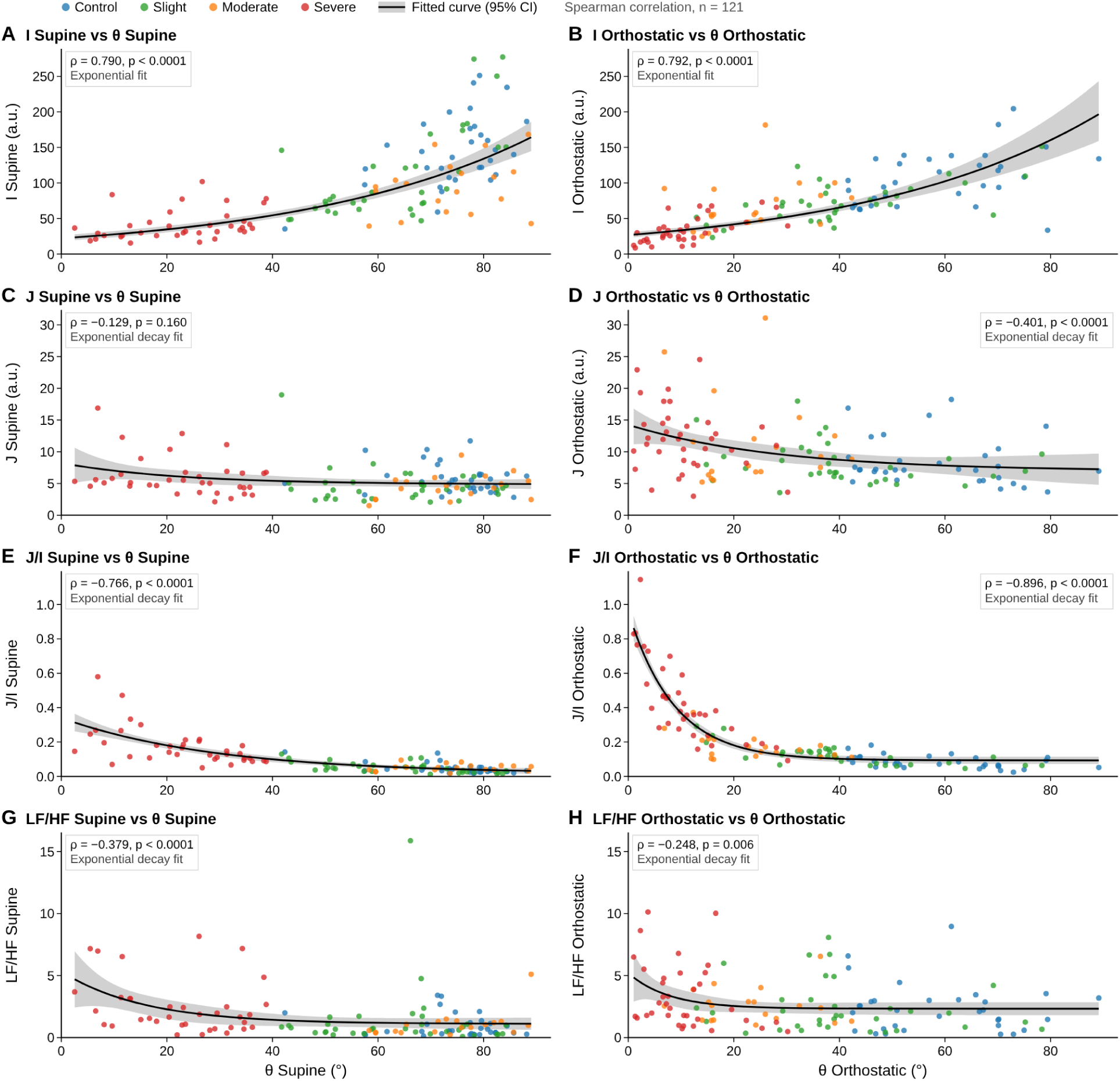
Relationship of wavelet-derived indices and conventional LF/HF ratio with θ BRS across postural conditions. (A,B) I, (C,D) J, (E,F) J/I, and (G,H) the conventional LF/HF ratio of IBI plotted against θ during supine (left) and orthostatic (right) periods. Each point represents one participant (n = 121), with colors indicating study group (Control, Slight, Moderate, and Severe). Black curves represent exponential fits with 95% confidence bands and are shown as visual guides. Spearman’s ρ and p values correspond to two-sided correlation tests. θ (°) represents the angle of the SBP–IBI ellipse, used as an estimate of baroreflex sensitivity. *I* and *J* are expressed in arbitrary units (a.u.), whereas J/I and LF/HF are dimensionless. The difference in absolute Spearman correlation coefficients (|ρ|) between J/I and LF/HF was assessed using bootstrap analysis.

***J/I*** was inversely associated with θ-derived BRS in both postures (Figure 5E, F), and the association was stronger during orthostasis (ρ = −0.896, p < 0.0001) than in the supine position (ρ = −0.766, p < 0.0001). The two postures differed in how ***J/I*** compared with ***I*** alone. In the supine position, where ***J*** was unrelated to θ-derived BRS, the association of ***J/I*** with θ-derived BRS was no stronger than that of ***I*** (|ρ| = 0.766 vs. 0.790). During orthostasis, where ***J*** was associated with θ-derived BRS, ***J/I*** showed a stronger association than ***I*** (|ρ| = 0.896 vs. 0.792).

The conventional LF/HF ratio was also inversely correlated with θ-derived BRS, although the associations were substantially weaker in both the supine position (ρ = −0.379, p < 0.0001) and during orthostasis (ρ = −0.248, p = 0.006; Figure 5G, H). In contrast to ***J/I***, the association between LF/HF and θ-derived BRS weakens during orthostasis. The difference in absolute correlation with θ between ***J/I*** and LF/HF was 0.39 in the supine position (95% bootstrap CI 0.26–0.52) and 0.65 during orthostasis (95% CI 0.50–0.81). Both intervals excluded zero, and none of the 10,000 bootstrap resamples favored LF/HF in either posture.

Overall, the association of ***J/I*** with θ-derived BRS was stronger during orthostasis, coinciding with the emergence of an inverse association between ***J*** and θ. This pattern suggested that ***J*** might contribute information about orthostatic θ beyond that captured by ***I*** alone. We therefore examined this possibility using nested regression models in the following section.

### 3.6 *J* adds information about orthostatic θ-derived BRS beyond *I*

Given that ***J*** was associated with θ-derived BRS more strongly during orthostasis (Section 3.5), we tested whether it contributed information about orthostatic θ-derived BRS beyond that contained in ***I***. We compared two nested linear models for *ln*(θ*_ort_*_ℎ*o*_), evaluated with 100 repetitions of shuffled 10-fold cross-validation (Figure 6). The reduced model included *ln*(*I_ort_*_ℎ*o*_) only:

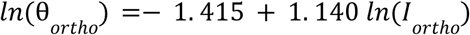

**Figure 6.**
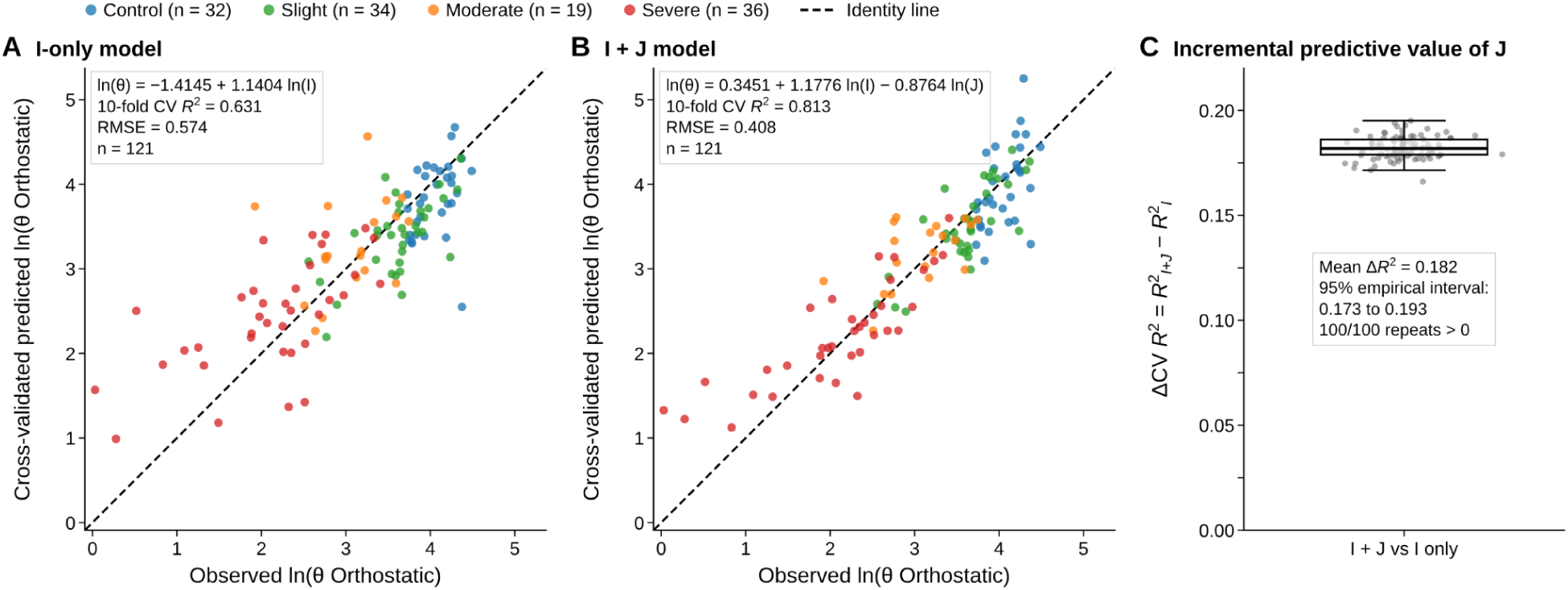
Contribution of *J* beyond *I* to the prediction of orthostatic θ. (A,B) Observed versus cross-validated predicted ln(θ) during orthostasis for the model including *I* alone (A) and the model including both *I* and *J* (B). Each point represents one participant (n = 121), with colors indicating study group (Control, Slight, Moderate, Severe). The dashed line indicates perfect agreement between observed and predicted values. Equations show the coefficients estimated from the full-sample models. R² and RMSE (log units) are reported as means across 100 repetitions of shuffled 10-fold cross-validation. (C) Gain cross-validated R² obtained by adding *J* to the model containing *I* (ΔR² = R²(I+J) − R²(I)) across the 100 repetitions. Each grey point represents one repetition, and the box plot summarizes the distribution. The 95% empirical interval reflects variability due to fold assignment rather than sampling uncertainty. All variables correspond to orthostatic measurements.

It explained a substantial part of the variance in orthostatic θ-derived BRS (mean cross-validated R² = 0.631; RMSE = 0.574 log units). Adding *ln* (*J_ort_*_ℎ*o*_) *ln*(θ*_ort_*_ℎ*o*_) = 0. 345 + 1. 178 *ln*(*I _ort_*_ℎ*o*_) − 0. 876 *ln* (*J_ort_*_ℎ*o*_) gave the full model:

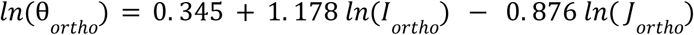

This increased the cross-validated R² to 0.813 and reduced the RMSE to 0.408 log units. The full model therefore roughly halved the unexplained variance (from 36.9% to 18.7%). On the original scale, the typical multiplicative prediction error decreased from about 1.78-fold to 1.50-fold.

The improvement was consistent across different assignments of participants to the cross-validation folds. The mean increase in cross-validated R² was 0.182 (2.5th–97.5^th^ percentile across repetitions, 0.173–0.193), and ΔR² was positive in all 100 repetitions (Figure 6C). The ***I***-only model systematically overestimated θ in participants with the lowest orthostatic values, most of whom belonged to the severe subgroup (Figure 6A). Including ***J*** reduced, although it did not eliminate, this bias and brought predictions closer to the identity line across the range of θ (Figure 6B).

In the full model, the coefficients of ln(***I***) and ln(***J***) had opposite signs and comparable magnitudes. Orthostatic θ-derived BRS was positively related to ***I*** and inversely related to ***J***, such that the model approximated a power function of the ratio ***J/I***. This is consistent with the strong association between ***J/I*** and θ-derived BRS described in Section 3.5. Taken together, these results suggest that, during orthostasis, the LF SBP information captured by ***J*** provides complementary information to the HF IBI information captured by ***I*** in accounting for the θ-defined baroreflex phenotype.

### 3.7 ROC-based evaluation of proposed and conventional indices for identifying θ-defined baroreflex impairment

Finally, we assessed how well each index identified θ-defined baroreflex impairment in each posture (Figure 7; Table 4). During orthostasis, J/I showed the highest discrimination of all indices (AUC 0.951, 95% CI 0.915–0.986), followed by ***I*** (0.885, 0.826–0.943) and HF power of IBI (0.864, 0.801–0.928) (Figure 7A). **J** alone showed moderate discrimination (0.705, 0.610–0.800), and the conventional LF/HF ratio was the lowest (0.623, 0.519–0.726). At the optimal cut-off, orthostatic J/I combined a sensitivity of 0.87 (0.77–0.93) with a specificity of 0.91 (0.79–0.96) and had the highest accuracy (0.88) and Youden index (0.78) of all indices evaluated (Table 4). Orthostatic ***I*** reached a higher specificity (0.96) and positive likelihood ratio (15.99), but at a lower sensitivity (0.71). LF/HF correctly identified fewer than one in three reference-positive participants (sensitivity 0.30).

**Figure 7.**
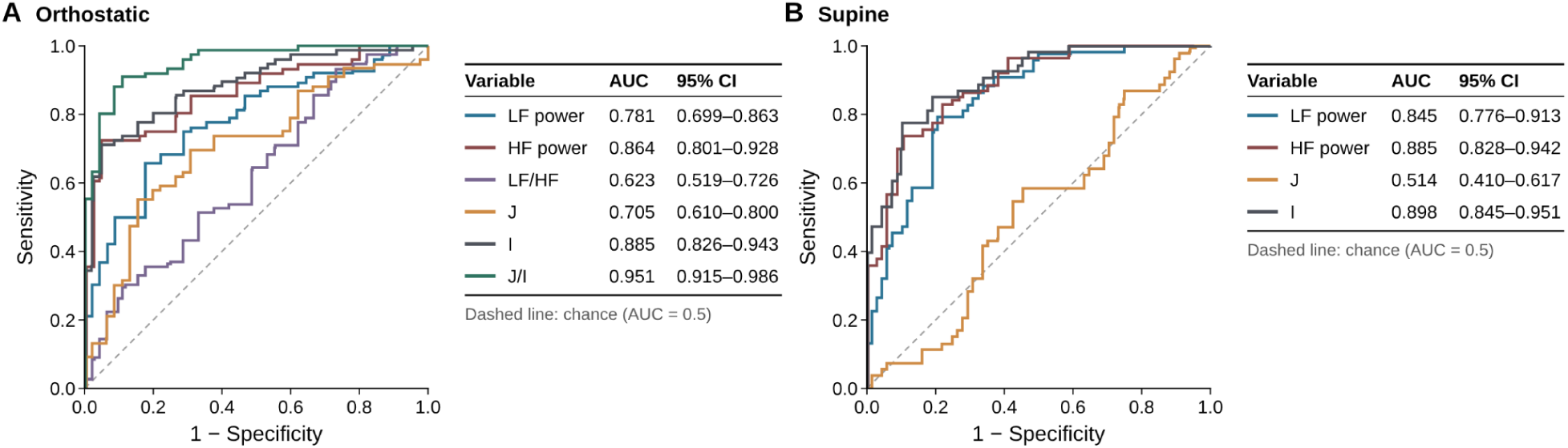
Receiver operating characteristic (ROC) analysis of conventional HRV and proposed wavelet indices for the classification of θ-defined baroreflex impairment. **(A)** Orthostatic and (B) Supine ROC curves for conventional HRV indices derived from IBI (LF power, HF power, and LF/HF ratio) and proposed wavelet-derived indices (I, J, and J/I). Baroreflex impairment was defined as θ < 68.3° in the supine position and θ < 40.3° during orthostasis. Lower values were considered test-positive for LF power, HF power, and I, whereas higher values were considered test-positive for LF/HF, J, and J/I. The dashed diagonal represents chance-level discrimination (AUC = 0.5). Abbreviations: AUC, area under the curve; HF, high frequency; HRV, heart rate variability; IBI, interbeat interval; LF, low frequency. (A) Supine and (B) orthostatic values of conventional HRV indices derived from IBI (LF power, HF power, and LF/HF ratio) and proposed wavelet-derived indices (I, J, and J/I).

**Table 4.** Diagnostic performance of conventional HRV and proposed wavelet indices for θ-defined baroreflex impairment. Values are presented as estimates with 95% confidence intervals (CIs) at the optimal cut-off for each index, determined by maximizing the Youden index. Lower values were considered test-positive for LF power, HF power, and I, whereas higher values were considered test-positive for LF/HF, J, and J/I. Cut-offs are expressed in ms² for LF and HF power, arbitrary units (a.u.) for *I* and J, and dimensionless units for LF/HF and J/I. CIs were calculated using the Wilson score method for proportions, the logarithmic method for likelihood ratios, and the Newcombe hybrid score method for the Youden index. Cut-offs were derived and evaluated in the same sample without external validation. Abbreviations: CI, confidence interval; HF, high frequency; HRV, heart rate variability; IBI, interbeat interval; LF, low frequency; LR+, positive likelihood ratio; LR−, negative likelihood ratio; NPV, negative predictive value; PPV, positive predictive value.

| **Variable** | **Sensitivity<br>(95% CI)** | **Specificity<br>(95% CI)** | **PPV<br>(95% CI)** | **NPV<br>(95% CI)** | **LR+<br>(95% CI)** | **LR−<br>(95% CI)** | **Accuracy<br>(95% CI)** | **Youden index<br>(95% CI)** |
|---|---|---|---|---|---|---|---|---|
| **Canonical indexes** |  |  |  |  |  |  |  |  |
| LF supine | 0.87<br>(0.77–0.93) | 0.57<br>(0.43–0.69) | 0.72<br>(0.61–0.81) | 0.77<br>(0.62–0.87) | 2.00<br>(1.45–2.76) | 0.23<br>(0.12–0.45) | 0.74<br>(0.65–0.81) | 0.43<br>(0.27–0.57) |
| LF orthostatism | 0.86<br>(0.76–0.92) | 0.53<br>(0.39–0.67) | 0.76<br>(0.66–0.83) | 0.69<br>(0.52–0.81) | 1.83<br>(1.32–2.54) | 0.27<br>(0.15–0.50) | 0.74<br>(0.65–0.81) | 0.39<br>(0.22–0.54) |
| HF supine | 0.78<br>(0.67–0.86) | 0.83<br>(0.71–0.91) | 0.85<br>(0.75–0.92) | 0.75<br>(0.62–0.84) | 4.59<br>(2.50–8.44) | 0.27<br>(0.17–0.42) | 0.80<br>(0.72–0.86) | 0.61<br>(0.44–0.72) |
| HF orthostatism | 0.72<br>(0.61–0.81) | 0.93<br>(0.82–0.98) | 0.95<br>(0.86–0.98) | 0.67<br>(0.54–0.77) | 10.86<br>(3.61–32.68) | 0.30<br>(0.20–0.43) | 0.80<br>(0.72–0.86) | 0.66<br>(0.50–0.76) |
| LF/HF orthostatism ratio | 0.30<br>(0.21–0.41) | 0.89<br>(0.77–0.95) | 0.82<br>(0.64–0.92) | 0.43<br>(0.33–0.53) | 2.72<br>(1.11–6.66) | 0.78<br>(0.65–0.94) | 0.52<br>(0.43–0.61) | 0.19<br>(0.04–0.32) |
| **Non-canonical indexes** |  |  |  |  |  |  |  |  |
| J supine | 0.37<br>(0.26–0.49) | 0.60<br>(0.47–0.72) | 0.54<br>(0.40–0.68) | 0.43<br>(0.32–0.54) | 0.93<br>(0.59–1.46) | 1.05<br>(0.79–1.39) | 0.47<br>(0.38–0.56) | −0.03<br>(−0.20–0.14) |
| J orthostatism | 0.68<br>(0.57–0.78) | 0.69<br>(0.54–0.80) | 0.79<br>(0.67–0.87) | 0.56<br>(0.43–0.69) | 2.20<br>(1.39–3.49) | 0.46<br>(0.31–0.67) | 0.69<br>(0.60–0.76) | 0.37<br>(0.19–0.52) |
| I supine | 0.81<br>(0.70–0.88) | 0.79<br>(0.67–0.88) | 0.83<br>(0.73–0.90) | 0.76<br>(0.64–0.86) | 3.90<br>(2.27–6.68) | 0.24<br>(0.15–0.40) | 0.80<br>(0.72–0.86) | 0.60<br>(0.43–0.72) |
| I orthostatism | 0.71<br>(0.60–0.80) | 0.96<br>(0.85–0.99) | 0.96<br>(0.88–0.99) | 0.66<br>(0.54–0.76) | 15.99<br>(4.09–62.43) | 0.30<br>(0.21–0.43) | 0.80<br>(0.72–0.86) | 0.67<br>(0.51–0.76) |
| J/I orthostatism ratio | 0.87<br>(0.77–0.93) | 0.91<br>(0.79–0.96) | 0.94<br>(0.86–0.98) | 0.80<br>(0.68–0.89) | 9.77<br>(3.82–25.00) | 0.14<br>(0.08–0.26) | 0.88<br>(0.82–0.93) | 0.78<br>(0.63–0.86) |

In the supine position***, I*** (AUC 0.898) and HF power (0.885) showed the highest discrimination, followed by LF power (0.845), whereas **J** did not discriminate since the vasoconstriction is only necessary after the orthostatic perturbation (0.514, 0.410–0.617; Figure 7B), consistent with the absence of an association between supine **J** and θ-derived BRS. As in the correlation and regression analyses, the advantage of combining **J** and ***I*** therefore emerged during orthostasis.

### 3.8 Illustrative case: window-of-interest analysis of a symptomatic period

Previous FFT-based analysis lacks time resolution for short period events; our proposed method is able to detect transient autonomic imbalance events. We applied the window-of-interest analysis as illustrated in Figure 8 in a patient from the severe subgroup who reported dizziness and headache during the period of orthostasis highlighted in blue (793–1002 s). During this symptomatic period, SBP repeatedly exceeded 170 mmHg, accompanied by concurrent increases in HR, and LF SBP power showed repeated elevations (Figures 8A, D). ***J*** computed over this window was 14.88, compared with 9.51 for the whole orthostatic segment and 2.14 for the supine segment, whereas ***I*** changed less (56.01 vs. 48.83 and 47.04, respectively). ***J/I*** was correspondingly higher in the window section (0.27) than over the whole orthostatic segment (0.19); this demonstrates that our method is able to detect transient episodes. The vasomotor episode was thus partly diluted when the whole orthostatic segment was averaged, highlighting the relevance of our approach.

**Figure 8.**
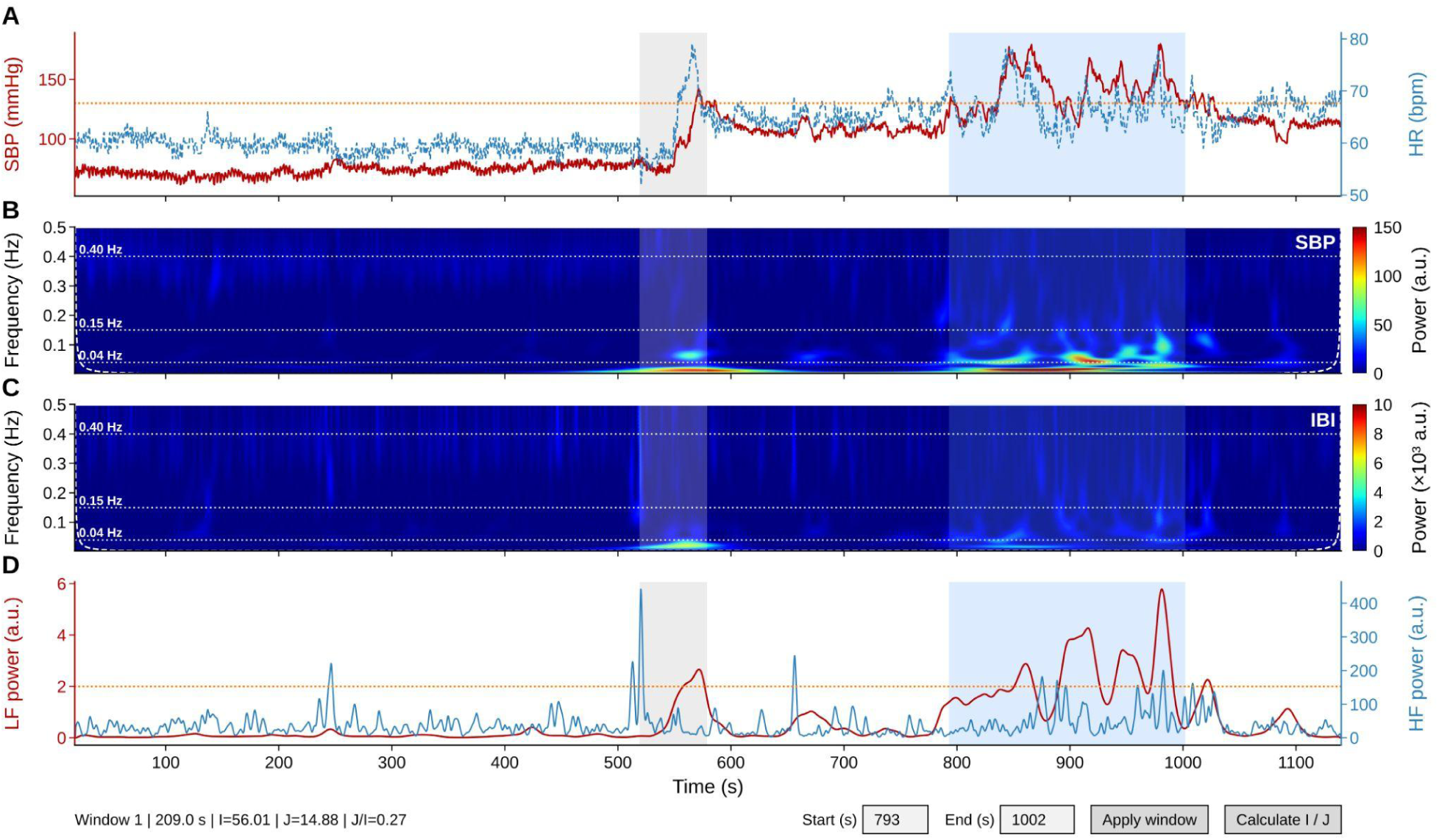
Wavelet detects sympathetic hyperactivity during a spontaneous vascular event. **(A)** Beat-to-beat systolic blood pressure (SBP, red) and heart rate (HR, blue) during the active standing test. (B, C) Wavelet power scalograms of SBP and IBI, respectively. Dotted lines indicate the frequency boundaries (0.04, 0.15, and 0.40 Hz), and the dashed contour marks the cone of influence (COI). (D) Time-varying low-frequency (LF) power of SBP (red) and high-frequency (HF) power of IBI (blue), as displayed by the analysis tool, which allows the user to select a time window and calculate the corresponding indices. In all panels, the grey band indicates the excluded posture-transition interval, and the blue band highlights the window of interest (793–1002 s; 209 s), during which the patient reported dizziness and headache. Orange dotted lines indicate visual reference levels used during clinical review (130 mmHg in A and 2 a.u. in D), at which SBP elevations and LF SBP power excursions were observed to coincide in this cohort. These reference levels do not represent the burst detection threshold, which is defined relative to the local baseline. During the symptomatic window, SBP repeatedly exceeded 170 mmHg, accompanied by increases in HR, while LF SBP power repeatedly exceeded 2 a.u.

## 4 Discussion

### 4.1 Principal findings

Given the recent SARS-CoV-2 pandemic, a controversy regarding if the infection affected autonomic balance arose. The canonical method was the LF/HF FFT-derived index, but this lacked temporal resolution and when it was averaged over a stationary period, important information was missed. We also identified that vasomotor sympathetic hyperactivity was not uniform over time; thus, we needed a method that allowed us to clearly identify sequences and patterns over time.

Then we introduced two indices derived from the continuous wavelet transform of routine beat-to-beat Finometer recordings: ***I***, from HF oscillations of IBI, and ***J***, based on LF oscillations of SBP and their transient elevations. We evaluated both indices, together with their ratio ***J/I***, against the θ-based baroreflex phenotypes in post-COVID-19 conditions and healthy controls.

Four main findings emerged. First, ***I*** closely reproduced conventional vagal HRV indices, whereas ***J*** was only weakly related to LF power of IBI, suggesting that it captured information on LF SBP dynamics not represented by IBI-based HRV. Second, ***I*** was reduced in all patient phenotypes in both postures, whereas the orthostatic increase in ***J*** was greatest in the subgroups with the lowest orthostatic θ-derived BRS. Consequently, ***J/I*** distinguished all patient phenotypes from controls during orthostasis, whereas the conventional LF/HF ratio did not. Third, ***J/I*** was strongly and inversely associated with θ at the individual level, particularly during orthostasis (ρ = −0.90), and more strongly than LF/HF in both postures. Fourth, ***J*** contributed information about orthostatic θ beyond ***I***, increasing the cross-validated explained variance from 63% to 81%.

To the best of our knowledge, this approach differs from previous wavelet-based methods in three respects: It characterizes cardiac and blood-pressure oscillations using their respective signals and frequency bands, explicitly incorporates transient elevations within a posture, and allows the indices to be recalculated over user-selected time windows.

In the context of post-COVID-19 dysautonomia, these findings are relevant because cardiovascular manifestations are heterogeneous, and a single measure of heart-rate variability may not capture the different patterns of cardiac and blood-pressure regulation. The present indices should therefore be viewed as complementary tools for physiological phenotyping, rather than as diagnostic tests or direct measures of specific autonomic pathways.

### 4.2 Index *I* as a time-resolved marker of cardiac vagal modulation

Index ***I*** was almost perfectly rank-correlated with the HF power of IBI (ρ = 0.97–0.98) and strongly correlated with RMSSD (ρ = 0.92–0.93) in both postures, across the full range from controls to severe subgroup participants. This agreement is expected because ***I*** and conventional HF power both quantify HF oscillations of IBI, which predominantly reflect respiratory sinus arrhythmia and cardiac vagal modulation (Malik et al., 1996). It also indicates that the wavelet pipeline preserves information captured by established vagal indices, despite the use of resampling, generalized Morse wavelet and COI masking. Index ***I*** can therefore serve as the cardiac reference against which ***J*** is contrasted. The added value of ***I*** lies less in its segment value than in its form. Because it is derived from a continuous band-power curve, it can be followed over time and recomputed over any window. Wavelet-based estimates of the same bands have detected autonomic changes that Fourier analysis missed or detected afterwards (Pichot et al., 1999; Wang et al., 2021).

At the group level, ***I*** was reduced in all three phenotypes in both postures. The reduction was most marked in the severe subgroup and paralleled that of HF power, RMSSD and pNN50, indicating that ***I*** added limited information for distinguishing phenotypes beyond conventional vagal indices. In the severe subgroup, the orthostatic decrease in ***I*** was much smaller than in controls (median Δ***I*** −7.1 vs. −42.8), which may reflect a floor effect because HF power was already markedly reduced in the supine position (Figure 1F). Index ***I*** was also strongly associated with θ-derived BRS (ρ ≈ 0.79 in both postures), consistent with the shared use of IBI in both measures. However, this association did not strengthen during standing. Thus, reduced cardiac vagal modulation was common across the post-COVID-19 phenotypes studied, whereas the between-phenotype information that emerged under orthostatic stress was more apparent in the SBP derived component ***J***.

### 4.3 Index *J* captures vasomotor oscillations and their transients, information absent from IBI-based HRV analysis

Index ***J*** is derived from LF oscillations of SBP, which include the approximately 0.1 Hz fluctuations commonly referred to as Mayer waves. These oscillations are associated with sympathetic vasomotor modulation and may increase during orthostatic or other sympathetic challenges (Julien, 2006). In healthy subjects, LF SBP can be temporally coupled to LF oscillations in muscle sympathetic nerve activity (MSNA) (Brychta et al., 2007). Orthostatic stress increases LF oscillations of MSNA together with those of arterial pressure (Furlan et al., 2000), and simulated orthostatic stress (lower-body negative pressure) strengthens the coupling between them (Cooke et al., 2009). Consistent with this physiology, ***J*** increased on standing in all groups. The novel observation was that the magnitude of this increase differed across phenotypes: orthostatic ***J*** and Δ***J*** were greatest in the subgroups with the lowest orthostatic θ-derived BRS, and ***J*** became associated with θ-derived BRS during orthostasis.

Two non-mutually exclusive mechanisms may contribute to this pattern. First, the LF SBP oscillations may themselves be larger because sympathetic vasomotor drive increases more on standing in patients with impaired baroreflex function. Second, the same oscillations may be less buffered. Previous work suggests that cardiac autonomic influences can modify Mayer-wave amplitude, indicating that heart rate oscillations normally buffer rather than reinforce them (Julien, 2006). Patients with low θ-derived BRS and low ***I*** appear to have lost much of this cardiac buffering, so a given vasomotor drive would be expected to produce larger SBP oscillations. Without direct sympathetic recordings, the present data cannot separate these mechanisms. Both, however, point to the same clinical picture: SBP is less well controlled when the baroreflex is challenged.

The weak association between ***J*** and LF power of IBI may reflect the fact that these measures characterize different aspects of cardiovascular regulation. At rest, LF oscillations of IBI are almost entirely the baroreflex-mediated response to sympathetically driven SBP oscillations, and both are abolished or markedly reduced when the vascular response to sympathetic drive is blocked (Cevese et al., 2001). The LF power of IBI therefore depends both on the input oscillation and on its transfer through the cardiac baroreflex. This explains why it tracks baroreflex function rather than only cardiac sympathetic tone and is low in autonomic failure (Goldstein et al., 2011). The pattern in the severe subgroup during orthostasis fits this interpretation. This subgroup had the highest ***J*** of all groups together with the lowest LF power of IBI, as expected when large SBP oscillations are no longer transmitted to the heart. An index computed from IBI alone sees only the output of the reflex. When transmission from SBP oscillations to IBI LF fails, the output can be small even while vasomotor oscillations are large, which may explain why LF/HF failed to separate the phenotypes during orthostasis. ***J*** reads the input side of the reflex directly.

The burst component of ***J*** addresses the observation that motivated this work: transient episodes within a single posture that may be obscured when each posture is averaged by a single value. Rapid rises in blood pressure during standing elicit transient baroreflex responses, and resolving such events requires a time resolution that long Fourier windows cannot provide (Mitsuyama et al., 2024). The CWT provides continuous time-frequency information, and the burst rate explicitly incorporates transient elevations in LF SBP power. The present analysis did not separate the contributions of burst magnitude and burst rate to group differences; the independent contribution of the burst component remains to be established. Accordingly, ***J*** is best interpreted as a composite marker of LF SBP oscillatory behavior, suitable for characterizing group differences and within-subject changes, rather than as an individual-level estimate of sympathetic nerve activity. Within-subject change is precisely how ***J*** distinguished the moderate subgroup from controls.

These findings may be relevant to cardiovascular dysautonomia in post-COVID-19 conditions, which has been associated with disturbances in orthostatic blood pressure and heart rate regulation. In this context, ***J*** may provide complementary information on vasomotor cardiovascular dynamics that is not captured by IBI-based HRV indices alone. By incorporating both the magnitude and temporal distribution of LF SBP oscillations, ***J*** may help characterize heterogeneous patterns of cardiovascular regulation during orthostatic stress and also provides a time window evaluation that allows a better inspection of vascular risk factors.

### 4.4 *J/I* tracks baroreflex function because it contrasts the regulated variable with its buffering response

***J/I*** was the index most closely associated with θ-derived BRS. The association was strong in the supine position (ρ = −0.77) and stronger during orthostasis (ρ = −0.90); in both postures it exceeded that of LF/HF by a margin whose bootstrap confidence interval excluded zero. The ratio form was also supported by the data. When *ln(**I**)* and *ln(**J**)* were entered separately into the model for orthostatic θ, their coefficients had opposite signs and comparable magnitudes, so the data themselves approximated a power function of ***J/I***. Adding ***J*** to ***I*** increased cross-validated explained variance by 18 percentage points, and this improvement was consistent across all cross-validation repetitions.

One possible interpretation is that ***J/I*** combines complementary features of cardiovascular regulation. Heart rate behaves as a physiological response whose variability reflects the capacity to buffer perturbations, whereas blood pressure is the regulated variable whose stability depends on that response. With aging and disease, the variability of the response decreases and that of the regulated variable increases (Fossion et al., 2018). ***J/I*** expresses this relationship. Its numerator quantifies LF oscillations of the regulated variable, SBP, and its denominator quantifies HF oscillations of the buffering response, IBI.

A failing baroreflex pushes both terms toward a higher J/I: less cardiac vagal modulation and larger, less-buffered vasomotor oscillations. The ratio therefore amplifies a pattern that each component shows only partially. This pattern aligns with the behavior experienced during each posture. In the supine position, where ***J*** was unrelated to θ-derived BRS, ***J/I*** was no more strongly associated with θ than ***I*** alone (|ρ| = 0.77 vs. 0.79). During orthostasis, when ***J*** became associated with θ, ***J/I*** showed a stronger association than *I* alone (|ρ| = 0.90 vs. 0.79). These findings suggest that ***J/I*** may be particularly informative under orthostatic stress, because it also incorporates burst SBP information in the formula, although its physiological specificity requires further exploration.

The same framework may help explain why LF/HF did not distinguish the groups during orthostasis. Both of its terms come from the buffering response alone. LF power of IBI largely reflects the baroreflex-mediated transfer of SBP oscillations rather than cardiac sympathetic tone (Cevese et al., 2001; Goldstein et al., 2011), and LF and HF power of IBI are themselves strongly correlated (Goldstein et al., 2011). When cardiac vagal and baroreflex function are globally impaired, both terms fall, and their ratio can remain within the normal range. This is consistent with the convergence of LF/HF across groups during orthostasis and with broader concerns about assigning sympathetic and parasympathetic activity to separate IB frequency bands (Clemson et al., 2022). In contrast, ***J/I*** combines information from two different cardiovascular signals: SBP and IBI.

***J/I*** also differ from time-frequency BRS estimates. The spectral α-index relates IBI and SBP power within the same frequency band (Chao-Ecija and Dawid-Milner, 2023), while our proposed method analyzes the component in two different frequency bands (LF and HF). Whereas cross-time-frequency methods may restrict estimates to regions with significant coherence and a phase consistent with baroreflex action (Orini et al., 2012; Kazimierska et al., 2019; de Boer and Karemaker, 2019). Both quantify the transfer through the cardiac limb of the reflex. ***J/I*** does not require coherence and instead contrasts different bands of the two effector signals. Because *A_HF_* dominates ***I***, and ***J*** reduces to 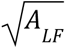 when there are no bursts detected, ***J/I*** approximates the square root of the ratio of LF SBP power to HF IBI power, but when there arebursts 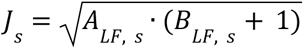 that incorporates the number of bursts and its activity pattern. In this context, ***J/I*** may help characterize patterns of baroreflex impairment that are not fully captured by cardiac variability indices alone, particularly during orthostatic stress. And its potential contribution to the identification of distinct autonomic phenotypes.

### 4.5 Implications across the θ-defined phenotypes

Autonomic dysfunction is frequent after SARS-CoV-2 infection (Goodman et al., 2021; Davis et al., 2023; Sasso et al., 2026). These manifestations are heterogeneous, and patients may differ in the extent to which cardiac and blood-pressure regulation are affected. Because the phenotypes in the present study were defined using the θ BRS indicator, our indices (***J***, ***I***) describe how the autonomic balance varies across groups of baroreflex function in this post-COVID-19 cohort.

In the severe subgroup, both limbs were affected in both postures. ***I*** was the lowest of all groups at rest and during orthostasis and changed relatively little on standing, consistent with an early loss of cardiac vagal modulation. This was the only subgroup in which ***J/I*** was already elevated in the supine position. The elevation was driven by the low ***I*** rather than by ***J***, which did not differ from controls at rest. On standing, these patients showed the largest increase in ***J*** and the highest orthostatic ***J/I***, together with the lowest LF power of IBI. This combination is consistent with large vasomotor oscillations that are no longer buffered by the heart. Thus, in this phenotype, differences were evident at rest and becomes more pronounced during the orthostatic challenge.

The moderate subgroup showed a different pattern. Supine BRS in this subgroup was similar to that of controls, with the difference that it fell markedly on standing. Correspondingly, ***J*** was normal at rest, and its absolute orthostatic value did not differ significantly from controls. However, the orthostatic change in ***J***, was approximately twice that of controls, and orthostatic ***J/I*** and Δ(***J/I***) were clearly elevated. This finding suggests the subgroup also had the largest increase in heart rate on standing. Its defining feature is thus a dynamic one: an exaggerated vasomotor response to standing, measurable only as a change from each patient’s own supine value. A single posture-averaged value, or an absolute threshold applied to orthostatic recordings alone, would have missed it. This highlights the importance of including both postures and expressing the orthostatic response as a within-subject change.

Slight, the subgroup that met neither θ-derived BRS criterion, had reduced ***I*** in both postures but no increased ***J***. Supine ***J*** was slightly lower than in controls, and orthostatic ***J*** and Δ***J*** did not differ. ***J/I*** was nevertheless elevated during orthostasis, mainly through the lower ***I***. This pattern suggests reduced cardiac vagally mediated modulation without a comparable increase in LF SBP power. ***J/I*** therefore captured a difference that was not apparent with LF/HF, although the clinical significance of this distinction requires evaluation in independent cohorts.

Two further observations support the view that ***J*** reflects the oscillatory component of blood pressure control rather than its mean level. First, the orthostatic change in SBP did not differ across groups, and the group medians remained below the 20 mmHg rise that defines an exaggerated orthostatic pressor response (Jordan et al., 2023), yet Δ***J*** differed. Second, across groups, orthostatic ***J/I*** followed the ordering of orthostatic θ-derived BRS (controls, slight, moderate, severe), and at the individual level it varied continuously with θ-derived BRS. Together with the strong individual-level association, this pattern suggests that ***J/I*** may be useful as a continuous marker of autonomic imbalance rather than only as a basis for categorical classification, in line with the spectrum concept proposed for θ-derived BRS (Sáinz-Jiménez et al., 2026).

### 4.6 Beyond segment averages: within-posture dynamics and window-of-interest analysis

This work was motivated by a clinical observation: during visual review of active standing recordings, transient episodes were noted within a single posture, including periods in which blood pressure and heart rate increased together. Such episodes disappear when each posture is summarized by a single spectral value. The case shown in Figure 8 illustrates the potential of the proposed method to examine these events. ***J*** computed over the window of interest was 14.88, compared with 9.51 for the whole orthostatic segment and 2.14 for the supine segment, whereas ***I*** changed comparatively little (56.01 vs. 48.83). Thus, averaging over the posture therefore diluted the vasomotor episode by about one third, while cardiac vagal modulation was essentially unchanged. This agrees with previous observations that wavelet analysis detects autonomic changes that averaged Fourier estimates miss or detect later (Pichot et al., 1999; Wang et al., 2021). The example is illustrative only: it is a single patient, and the window was chosen with knowledge of the reported symptoms.

Existing time-frequency approaches address related but different questions. Time-varying HRV indices follow LF and HF power of IBI over time (Pichot et al., 1999; Chen, 2002; Lu et al., 2005; Wang et al., 2021), but rely on IBI alone. Time-varying BRS methods estimate the transfer from SBP to IBI and may restrict estimates to time-frequency regions with significant coherence and a phase consistent with baroreflex action (Orini et al., 2012; Kazimierska et al., 2019; de Boer and Karemaker, 2019; Chao-Ecija and Dawid-Milner, 2023).

These approaches provide information about coupling between signals, but BRS may be difficult to estimate when significant coupling is absent, as reported in some populations with autonomic dysfunction or patients with diabetic autonomic neuropathy and heart transplant recipients, in whom no significant LF coupling was found (Orini et al., 2012). ***I*** and ***J*** instead characterize oscillatory features of each signal without requiring significant coupling. They describe the state of each effector even when the transfer between them is too weak to estimate, which is precisely the situation of the most impaired patients. Methods based on phase coherence and coupling between cardiovascular and respiratory oscillations provide further detail (Clemson et al., 2022; Miranda Hurtado et al., 2023). However, they generally require additional signals such as respiration or MSNA, together with surrogate testing, which limits their use in routine review. We therefore see the proposed indices as complementary to these methods rather than as substitutes for them.

From a practical standpoint, the method requires only the beat-to-beat SBP and pulse interval routinely obtained with a finger-cuff device during an active standing test. It does not require ECG, respiratory or nerve recordings for calculation of the proposed indices. The scalograms and band-power curves can be inspected alongside SBP and HR, and the indices can be recomputed over any window of at least 90 s identified during that inspection. The computational cost of the CWT is no longer a barrier: fast implementations allow analysis in real time (Arts and van den Broek, 2022), so the approach could in principle be applied during the test itself rather than only afterwards. However, clinical applications will require prospective validation, reproducibility testing and evaluation of symptom blinded window selection.

Overall, the window-of-interest approach extends cardiovascular assessment beyond segment-averaged measures by allowing transient changes in blood pressure and cardiac variability to be examined within a given posture. This may be particularly relevant to post-COVID-19 dysautonomia, where heterogeneous orthostatic cardiovascular responses (Goodman et al., 2021; Azcue et al., 2023) may not be fully represented by measurements averaged over an entire standing period.

Although further validation is needed, combining time-varying SBP oscillations with the proposed indices may provide a complementary approach to characterizing transient patterns of cardiovascular dysregulation and their potential relationship with orthostatic symptoms.

## 5. Limitations

This was a single-center, cross-sectional study with a modest sample size (n = 121) and no independent external validation cohort. Sex was not included as a covariate. Thus, the findings characterize the proposed indices (***J***, ***I***) across θ-derived BRS phenotypes but do not constitute independent diagnostic validation.

The physiological interpretation of ***J*** is also limited by the indirect nature of LF SBP power as a marker of vascular sympathetic modulation and lack of direct sympathetic recordings.

Methodologically, wavelet power was not normalized because the magnitude of the oscillations was relevant to seeing the correlation of the SBP magnitude. The burst threshold was defined for this specific cohort, and the magnitude and burst components of ***J*** were not examined separately. IBI was derived from the arterial pressure pulse rather than the ECG, and the sensitivity of the wavelet parameters to alternative settings remains to be assessed. Larger, independent studies incorporating direct sympathetic recordings, independent BRS measurements, respiratory monitoring, and standardized analytical procedures are needed to establish the reproducibility, physiological interpretation, and generalizability of these findings.

## 6. Future directions

Conventional HRV has been used as an autonomic marker in ischemic stroke (Lakusić et al., 2003) and critical illness (Winchell and Hoyt, 1996), and wavelet analysis of HRV has been applied during myocardial reperfusion (Toledo et al., 2003) and in hypertension (Takagi et al., 2006). These approaches rely on IBI alone, and SBP-derived wavelet variables have not yet been evaluated in clinical populations. The proposed variables could be tested in other conditions with impaired baroreflex function, such as diabetes mellitus, in which spontaneous BRS detects autonomic dysfunction earlier than conventional autonomic tests (Frattola et al., 1997). Longitudinal recordings in post-COVID-19 patients could determine whether variable ***J*** and ***J/I*** follow the fluctuating course of their symptoms (Owen et al., 2025), and repeated measurements could monitor the response to treatment. Finally, given the non-invasive nature of the proposed method, it could be implemented in wearable devices as a tool to estimate autonomic imbalance once cuffless beat-to-beat blood pressure measurement is sufficiently accurate (Qin et al., 2022), as wearable HRV monitoring is already feasible in long COVID (Ruijgt et al., 2026), contributing to preemptive medicine.

## 7. Conclusions

Wavelet-derived indices obtained from routine beat-to-beat recordings provide complementary, time-resolved measures of cardiac vagal modulation and low-frequency systolic blood pressure oscillations. The index ***I*** captured the expected patterns of cardiac vagal modulation, whereas ***J*** characterized LF SBP oscillations and their transient increases, providing information not captured by IBI-based HRV.

Their ratio (***J/I***) distinguished all θ-defined baroreflex phenotypes from controls during orthostasis and showed a stronger association with θ BRS estimation than the conventional LF/HF ratio; this leads us to conclude that ***J/I*** reflects more accurately the autonomic imbalance present in patients with different degrees of BRS impairment. Thus, ***J*** contributes information beyond ***I***. The visual representation of these indices and the window-of-interest analysis further illustrate their potential to identify transient within-posture dynamics that may be overlooked by segment-averaged approaches.

Together, these findings suggest that combining cardiac and vasomotor-related oscillatory information may complement conventional HRV and established BRS methods in the assessment of dysautonomia. This approach provides a framework for a more comprehensive characterization of autonomic cardiovascular regulation, with potential applications in identifying distinct hemodynamic patterns and transient physiological responses during orthostasis.

## Data Availability

Data is available under request.

