## Supplementary methods 1 for "Wavelet-derived indices of cardiac vagal and vasomotor oscillations as a new tool to better understand autonomic imbalance in post-COVID-19 dysautonomia"

Supplementary Methods S1

Supplementary Methods S1. Wavelet time-frequency analysis: choice of transform and parameter settings

Choice of transform. We used the CWT rather than the short-time Fourier transform (STFT) or the discrete wavelet transform (DWT). Unlike the STFT, the CWT provides a scale-dependent time-frequency representation, adapting its effective temporal resolution to the frequency being analyzed (Torrence and Compo, 1998; Mallat, 1999). Furthermore, the CWT allows power to be integrated over frequency bands with boundaries that can be specified to match the physiological ranges of interest. In contrast, conventional dyadic DWT decompositions divide the signal into frequency bands determined by successive decomposition levels, which may not align precisely with the predefined physiological band limits (Pichot et al., 1999; Chen, 2002).

Resampling and edge effects. PCHIP preserves the monotonicity of the data between successive beats while minimizing the overshoot of cubic spline interpolation (Fritsch and Carlson, 1980). Computing the CWT over the full 14-min recording avoided introducing an artificial signal boundary at the postural transition, although wavelet edge effects at the beginning and end of the full recording remained possible; these were limited by setting coefficients outside the COI to zero. The COI was defined from the time-domain width of the wavelet at each scale (Torrence and Compo, 1998).

Wavelet parameters. The generalized Morse wavelet family has two parameters: γ, which controls the symmetry of the wavelet, and the time-bandwidth product P² = βγ, which sets the trade-off between temporal and frequency resolution (Olhede and Walden, 2002; Lilly and Olhede, 2012). Generalized Morse wavelets were also used in previous wavelet-based baroreflex analyses (de Boer and Karemaker, 2019). Both signals were analyzed with γ = 3, which provides an approximately symmetric analytic wavelet with minimal time and frequency skewness (Lilly and Olhede, 2009). For SBP, P² = 20 (β = 20/3), favoring frequency resolution for the slower LF oscillations; for IBI, P² = 10 (β = 10/3), favoring temporal resolution for the faster and more transient HF oscillations. Both values were selected during preliminary calibration of the method. The analysis used 24 voices per octave over a frequency range of 0.003–1 Hz, ensuring that the frequency bands of interest were positioned well within the limits of the analyzed scale range.

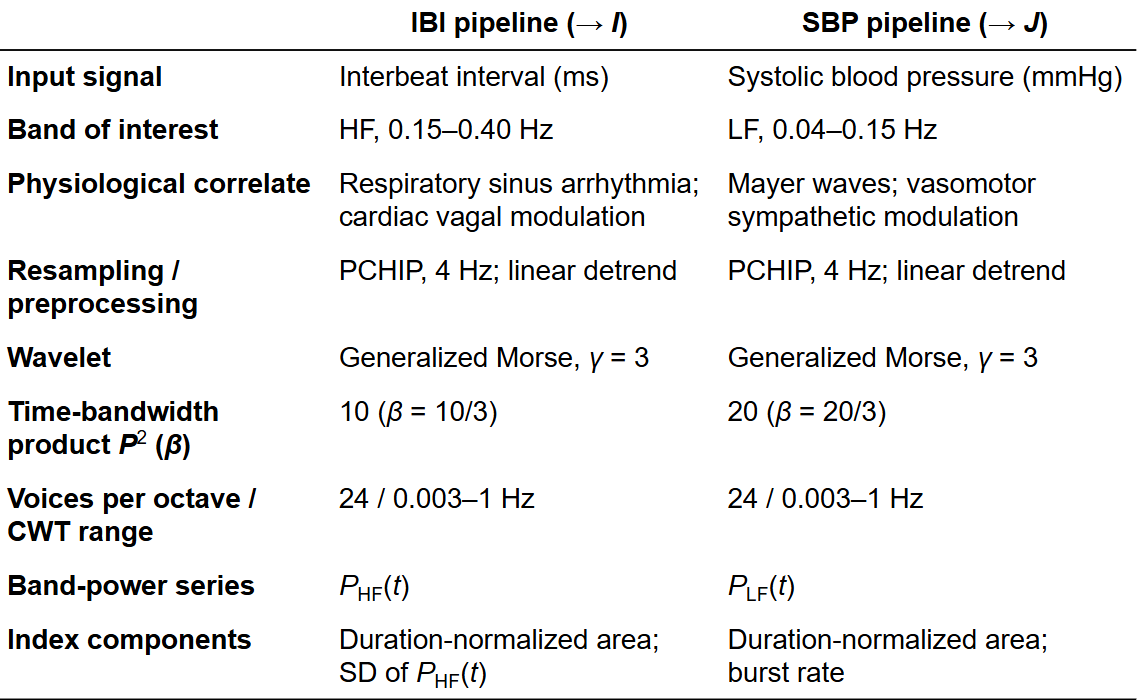

*Supplementary table 1. Wavelet analysis settings for the IBI and SBP pipelines.*

*Both signals were processed identically except for the band of interest and the time-bandwidth product. The CWT was computed with generalized Morse wavelets in ssqueezepy (version 0.6.5) over the entire recording. Coefficients outside the cone of influence were set to zero before band power was integrated with the trapezoidal rule (Section 2.4). The time-bandwidth product is P² = βγ. The physiological correlates indicate the predominant influence on each band and do not imply that band power directly measures autonomic nerve activity. Abbreviations: CWT, continuous wavelet transform; HF, high frequency; IBI, interbeat interval; LF, low frequency*

Wavelet power. Wavelet power was computed as the squared magnitude of the CWT coefficients, P(f,t) = |W_X(f,t)|², and coefficients outside the COI were set to zero, P_COI(f,t) = P(f,t) · 𝟙{f ≥ f_COI(t)}, before band power was integrated.
